# Coupling models of within-human, human-to-mosquito, and within-mosquito parasite dynamics to identify key drivers of malaria transmission

**DOI:** 10.1101/2025.10.28.25339021

**Authors:** Xiao Sun, Matthew W. A. Dixon, James S. McCarthy, James M. McCaw, Pengxing Cao

## Abstract

The transmission of malaria parasites from humans to mosquitoes is essential for the parasite’s life cycle. However, human-to-mosquito transmission remains poorly characterized due to insufficient quantification of biological parameters and limited tools for systematic study. To fill this knowledge gap, a mathematical model was fitted to data from multiple mosquito feeding experiments to estimate poorly quantified biological parameters. Drawing on these quantitative results, we developed a multi-scale model of human-to-mosquito transmission to identify the dominant host factors determining a human’s infectiousness to mosquitoes. Our primary findings are that, for individuals with asymptomatic infections, 1) the time from exposure to the onset of infectiousness is primarily driven by in-human parameters governing asexual parasite multiplication, sexual conversion, and gametocyte maturation; and 2) for those with established infection, infectiousness is predominantly influenced by the availability of male gametocytes in the human bloodstream, mosquito blood meal volume, and gamete fertilization efficiency within the mosquito midgut.

## Introduction

Malaria infection is a vector-borne disease that continues to pose a health threat to billions of people worldwide, causing approximately 610,000 deaths in 2024 [1]. The transmission of malaria parasites from the human host to a female Anopheles mosquito requires the mosquito to feed on the blood of the infected individual and ingest at least one male and one female mature gametocyte [2]. Those gametocytes undergo rapid transition into gametes, with each male gametocyte producing eight microgametes. Male microgametes and female macrogametes then undergo meiosis in the mosquito midgut to form diploid ookinetes that invade the midgut wall and divide to form oocysts. Each oocyst matures to release thousands of haploid sporozoites which move to the salivary gland of the mosquito, ready for transmission back to a human to cause a new infection, thus sustaining the parasite population. This process can be investigated experimentally by performing either direct feeding assays (DFA) where mosquitoes are allowed to feed on the skin of an infected individual, or by standard membrane feeding assays (SMFA) where infected blood is placed in a special feeding container for mosquitoes to feed on. Mosquitoes can be examined for the presence of oocysts or sporozoites; the proportion of mosquitoes developing oocysts or sporozoites provides a direct measurement of the probability that a mosquito will become infected or infectious after taking a blood meal from the infected human (referred to as the probability of human-to-mosquito transmission). Such assays are widely used to experimentally evaluate both the infectiousness of infected human hosts to mosquitoes and the efficacy of interventions, such as drugs or vaccines to block transmission [3–5].

For falciparum malaria, a quantitative understanding of the process of parasite transmission from a human to a mosquito remains incomplete, despite the availability of substantial mosquito feeding assay data collected over many years. This gap can be attributed to two main factors. The first is the incomplete quantification of key biological parameters that influence humanto-mosquito transmission. In particular, two biological parameters that modulate gamete development and fertilization, and hence influence transmission, have not yet been rigorously estimated. One is the ratio of viable male gametes produced in the mosquito midgut to male gametocytes taken in a blood meal (hereafter referred to as the male gamete-to-gametocyte ratio). Theoretically, this ratio has an upper bound of eight, as each male gametocyte produces up to eight male gametes [6]. However, the true value is expected to be substantially lower, since not all male gametes remain viable during replication and differentiation and therefore are incapable of successful fertilization [7–9]. The other parameter is the probability of a successful fertilization event between viable male and female gametes. The absence of estimates for the probability of fertilization in the literature is likely attributable to several limitations, including the lack of experimental studies quantifying all key parasite populations (e.g., male and female gametes and zygotes/oocysts), as well as the scarcity of modeling studies specifically designed to characterize the fertilization process.

The second factor contributing to our incomplete understanding of human-to-mosquito transmission is the lack of tools designed to quantitatively assess the effects of various host (human and mosquito) factors on the probability of human-to-mosquito transmission. A direct approach to overcome this limitation would be to design and conduct clinical, experimental or observational studies to collect such data. In previous studies, the focus has predominantly been on how the level of gametocytemia in human hosts influences the transmission probability. As illustrated in Fig. 1, where mosquito feeding assay data from three representative experimental studies are presented [10–12], there remains substantial unexplained variability in the probability of transmission when gametocytemia is considered the only predictor. The variability is particularly evident in asymptomatic infections, where transmission outcomes are highly heterogeneous across individuals and not solely determined by gametocyte density [13]. This has significant public health implications for implementing strategies to interrupt malaria transmission and highlights the importance of understanding the main drivers of human-to-mosquito transmission. To understand these drivers, we can bring together experimental data and mathematical models that capture the established (and hypothesized) mechanisms underlying key biological processes in human-to-mosquito transmission; that is gametocyte development in humans, parasite uptake during mosquito feeding, and parasite development within mosquitoes. Although numerous models have been developed to study within-host malaria dynamics and transmission, no comprehensive model of human-to-mosquito transmission has been developed. Existing models that examine aspects of human-to-mosquito transmission either lack within-mosquito dynamics [14–19], omit blood-stage dynamics [20–24], or fail to mechanistically capture the underlying biological processes such as gamete development and fertilization [25, 26].

**Figure 1:**
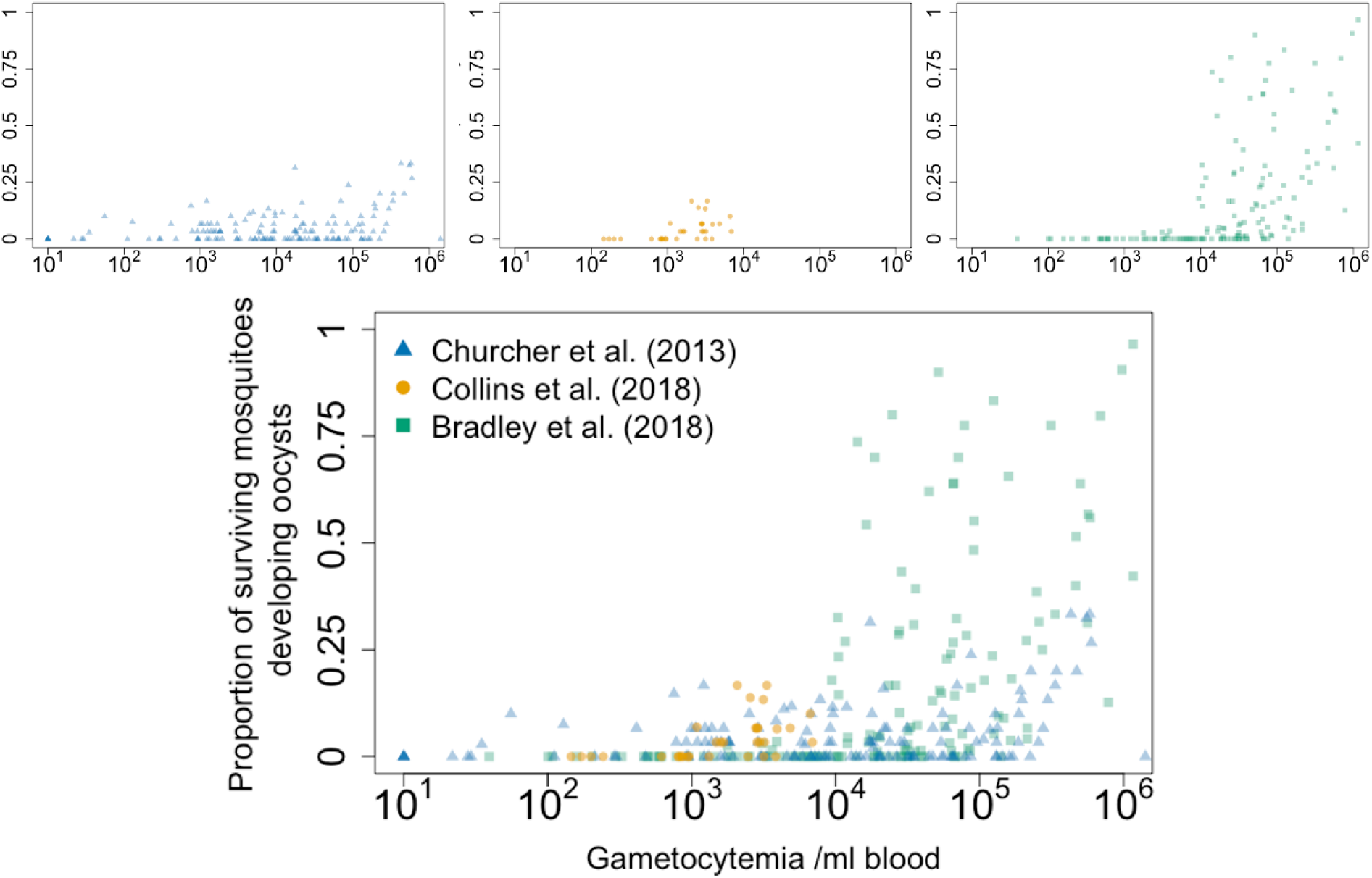
Direct feeding assay data from three representative experimental studies. Each data point represents a sample of gametocytemia collected from an infected human and the proportion of surviving mosquitoes that developed oocysts following a blood meal from the infected human and subsequently developed oocysts. Blue triangles represent data from Churcher et al. [10]. Yellow dots represent data from Collins et al. [11]. Green squares represent data from Bradley et al. [12].

Here, we develop and combine two mathematical models to estimate key biological parameters and investigate the relative importance of various host factors—including those governing gametocyte development in humans and those driving parasite’s development in mosquitoes— that determine the probability of human-to-mosquito transmission. The first model captures both gametocyte uptake by mosquitoes and parasite development within mosquitoes to describe mosquito feeding (hereafter referred to as the mosquito feeding model). The mosquito feeding model was fitted to mosquito feeding assay data from a volunteer infection study (VIS) by Collins et al. and a field study comprising sites across three countries by Bradley et al. (reproduced in Fig. 1), which provide measurements of sex-specific gametocyte densities and the corresponding probability of mosquito infection under different conditions (mosquito species and transmission settings). These data enabled us to estimate two poorly quantified biological parameters. Drawing on these estimates, we then developed a second model (hereafter referred to as the human-to-mosquito transmission model), which integrates the mosquito feeding model with an existing data-calibrated within-human parasite dynamics model [18,19]. This integrated model incorporates key host factors governing the full process of human-to-mosquito transmission, from asexual blood-stage expansion in humans to sporozoite formation in mosquitoes. The human-to-mosquito transmission model was used to characterize the probability of humanto-mosquito transmission in asymptomatic infections, and identify the key factors (i.e., model parameters) that influence this probability by conducting a global sensitivity analysis.

## Results

### Estimation of key biological parameters driving gamete development and fertilization

The male gamete-to-gametocyte ratio (*ρ*) and the probability of successful fertilization between a viable male gamete and a viable female gamete (*p*_*f*_) are key biological parameters influencing human-to-mosquito transmission of malaria, yet are poorly characterized in the literature. To estimate these parameters while also accounting for potential variability across different transmission settings, the mosquito feeding model was fitted separately to four mosquito feeding assay datasets: one dataset from a VIS conducted in Australia (reported by Collins et al. [11]) and three field datasets from Burkina Faso, Cameroon, and Mali (reported by Bradley et al. [12])). The locations, numbers of data points, and mosquito species associated with the four datasets are summarized in Table 1. The data used for model fitting include the measurements of male and female gametocyte concentration for each participant and the proportion of mosquitoes developing oocysts associated with each pair of gametocytemia measurements. The mosquito feeding model is a set of mathematical equations and statistical distributions that parameterize the turnover dynamics of parasites of different developmental stages from gametocytes to oocysts and sporozoites. Model fitting was conducted using an approximate Bayesian computation (ABC) algorithm to sample the parameter values with which the solutions to the mosquito feeding model match the longitudinal measurements based on a metric for goodness of fit. Details on the data, the model, the fitting algorithm, and statistical diagnostics are provided in the Materials and Methods.

**Table 1:** Summary of the data used in model fitting and parameter estimation results.

| Country | Mosquito species | MAP estimate of $\rho$<br>(95% HDPI) | MAP estimate of $p_f$<br>(95% HDPI) |
| --- | --- | --- | --- |
| Australia ($n = 33$) | *Anopheles stephensi* [11] | 0.796 (0.129–2.903) | 0.0298 (0.0059–0.1090) |
| Burkina Faso ($n = 64$) | *Anopheles coluzzii* [12] | 0.194 (0.015–1.531) | 0.0560 (0.0014–0.4269) |
| Cameroon ($n = 13$) | *Anopheles coluzzii* [12] | 0.274 (0.008–1.754) | 0.0130 (0.0004–0.1044) |
| Mali ($n = 71$) | *Anopheles coluzzii*, *Anopheles gambiae* s.s., and hybrid forms [12] | 0.239 (0.014–1.526) | 0.0144 (0.0004–0.1069) |

Note: $n$ is the sample size for each mosquito feeding study. Data from Australia were obtained from a VIS conducted by Collins et al. [11], in which mosquito feeding assays were conducted for 11 participants at three post-inoculation time points, yielding a total of 33 data samples. Detailed male and female gametocytemia data for each participant at each feeding time point are provided in S1 Fig. The corresponding proportions of mosquitoes developing oocysts, together with the model fitting results, are shown in S2 Fig. Data from Burkina Faso, Cameroon, and Mali were obtained from Bradley et al. [12], in which each data point corresponded to one mosquito feeding assay conducted using a blood sample from a single participant.

The model fitting results for the VIS dataset conducted in Australia and the three field datasets from Burkina Faso, Cameroon, and Mali are shown in Fig. 2. The model posterior predictive distributions of the proportion of surviving mosquitoes developing oocysts (violin plots in Panels a–d), ordered by ascending male gametocyte concentration, encompass the majority of observations, indicating a good fit to the experimental data. These results also show a consistent trend of an increasing proportion of infected mosquitoes with increasing male gametocyte concentration. Alternative visualizations of the fitting results, ordered by ascending female gametocyte concentration are provided in S3 Fig, which similarly show an increasing proportion of infected mosquitoes with increasing female gametocyte concentration.

**Figure 2:**
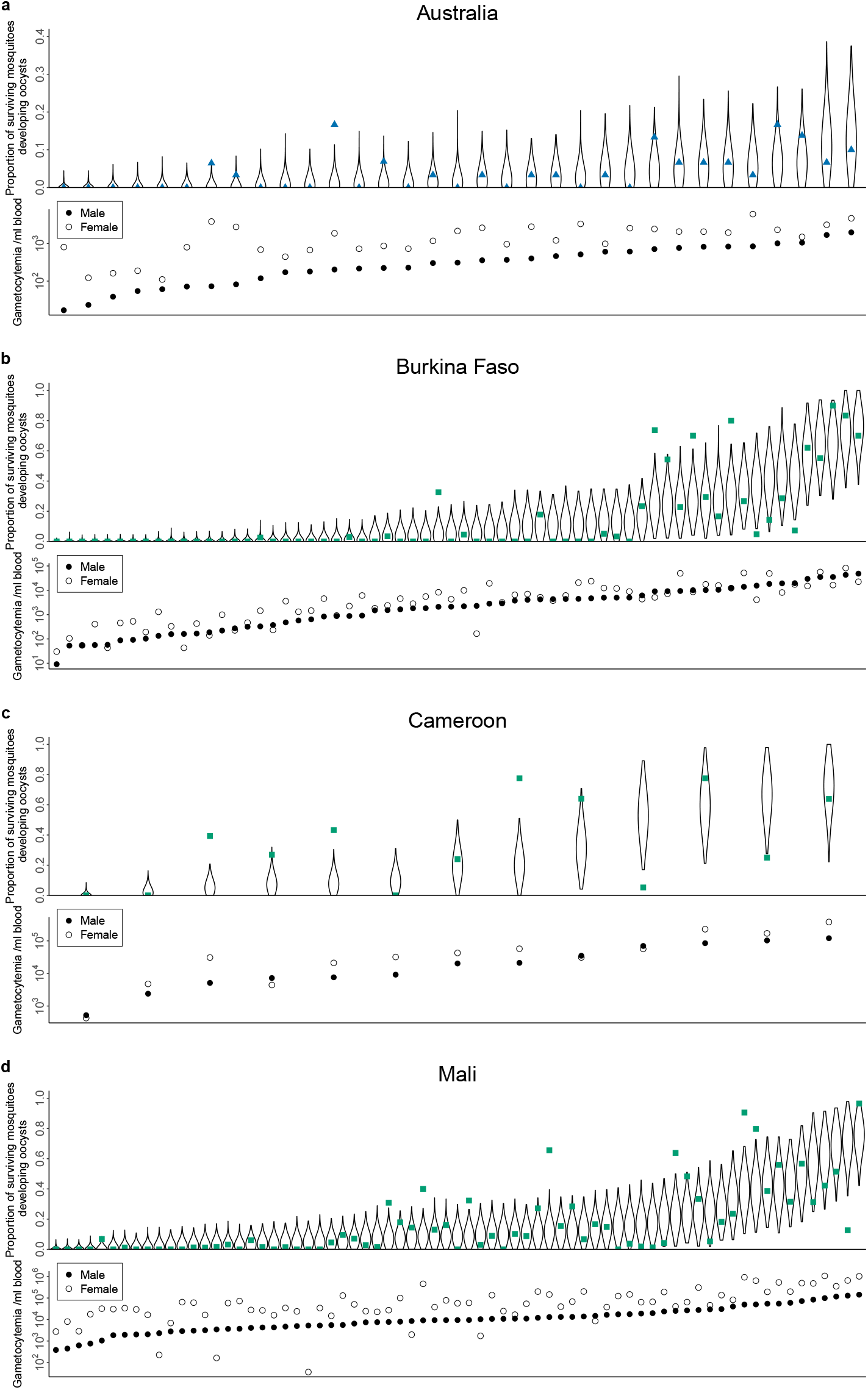
Results of model fitting. The model posterior estimates of the proportion of surviving mosquitoes developing oocysts for each data sample, ordered by male gametocytemia are shown for (a) Australia, (b) Burkina Faso, (c) Cameroon, and (d) Mali, respectively. Within each panel, samples are ordered by increasing male gametocytemia. Violin plots represent the distributions of the estimated proportions of mosquitoes developing oocysts corresponding to the 5000 accepted posterior samples obtained during model fitting. Blue triangles and green squares represent the experimental observations from Collins et al. [11] and Bradley et al. [12], respectively. Black dots and circles represent male and female gametocytemia, respectively. Note that the upper limit of proportion of surviving mosquitoes developing oocysts is set to 0.4 for the Australia (VIS) panel to illustrate the model estimates and experimental observations more clearly, whereas an upper limit of 1 is used for the other panels.

The samples drawn from the joint posterior distributions of *ρ* and *p*_*f*_ using the ABC algorithm are shown in Fig. 3 and summarized in Table 1 which reports the maximum a posteriori (MAP) estimate, defined as the point estimates corresponding to the highest density in the posterior distribution, and the estimated 95% highest density posterior interval (95% HDPI), which contains approximately 95% of the samples in the posterior distribution. For the male gamete-to-gametocyte ratio *ρ*, the MAP estimates varied across the four datasets. The estimate is highest for the VIS dataset conducted in Australia at 0.796 (95% HDPI: 0.129–2.903), followed among the three field datasets by Cameroon at 0.274 (0.008–1.754), Mali at 0.239 (0.014–1.526), and Burkina Faso at 0.194 (0.015–1.531). These MAP estimates of *ρ* suggest that every ten male gametocytes taken up in a mosquito blood meal lead to the formation of, on average, approximately two to eight viable male gametes available for fertilization in the mosquito midgut (implying an attrition rate of 90%–98% from the theoretical maximum of eight male gametes arising from one male gametocyte). The MAP estimates for the probability of successful fertilization per gamete pair, *p*_*f*_, are 0.0298 (95% HDPI: 0.0059–0.1090) for the VIS dataset conducted in Australia, and 0.0560 (0.0014–0.4269), 0.0130 (0.0004–0.1044), and 0.0144 (0.0004–0.1069) for the field datasets from Burkina Faso, Cameroon, and Mali, respectively. The results suggest that the two estimated parameters may vary significantly under different conditions, such as geographic area, transmission setting, and mosquito species. While application of our model to these four data sets has provided estimates of these two important biological parameters for the first time, we note the presence of residual correlation in the posterior distribution (i.e., the hyperbolic correlation in the joint posterior visible, albeit at low density, in Fig. 3). Substantially different parameter combinations (albeit within relatively low density regions)—high *ρ* with low *p*_*f*_ or conversely low *ρ* with high *p*_*f*_ —produce comparable measures of goodness of fit. We elaborate on the implications of this correlation in the Discussion.

**Figure 3:**
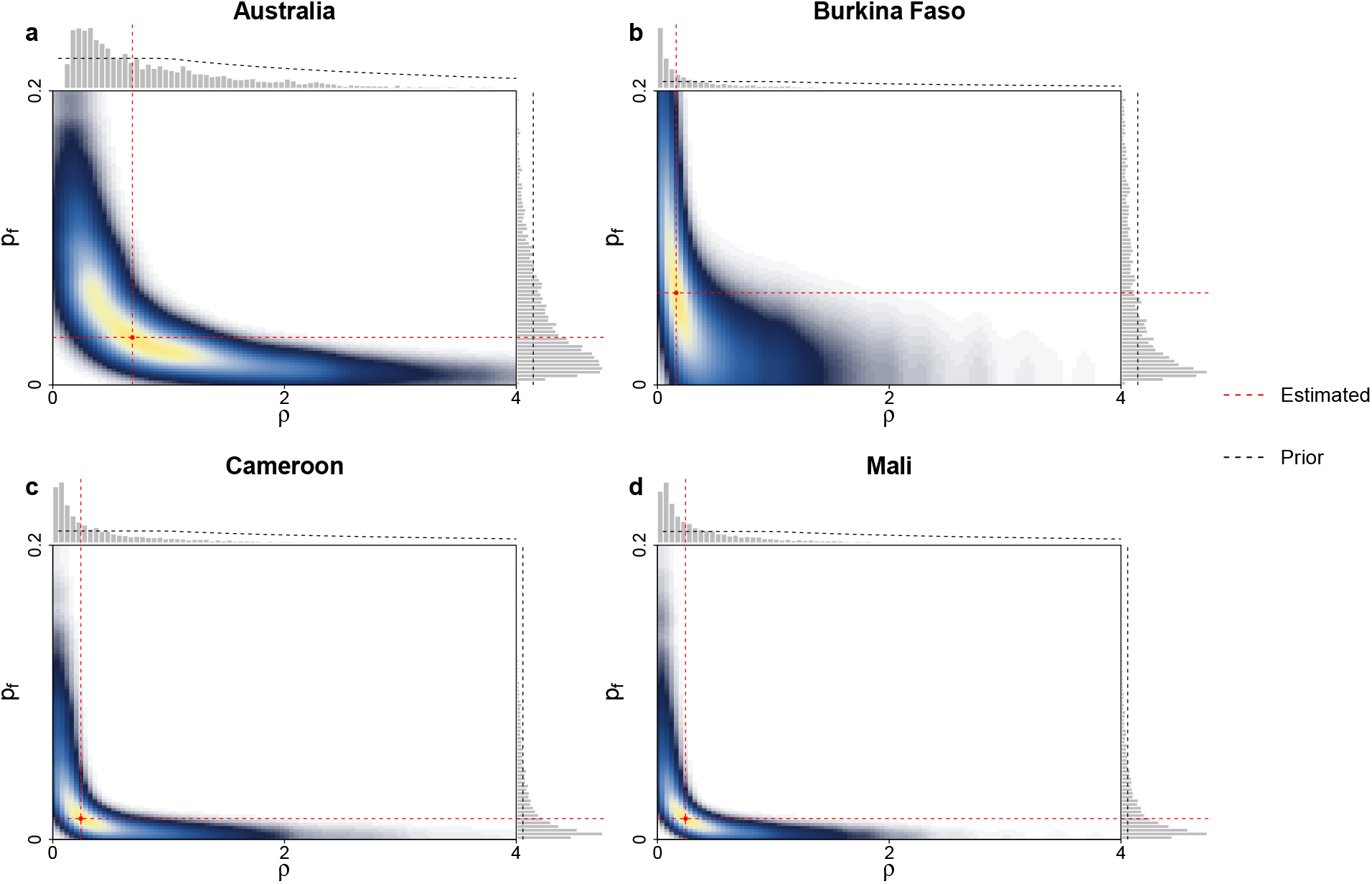
Results of parameter estimation. The joint and marginal distributions of 5000 draws from the posterior (i.e., estimate) for the male gamete-to-gametocyte ratio (*ρ*) and the probability of successful fertilization per gamete pair (*p*_*f*_) are shown for (a) Australia, (b) Burkina Faso, (c) Cameroon, and (d) Mali. Black dashed curves indicate the prior distributions of the parameters. The red dashed lines intersect at the MAP estimates (the point with the highest posterior density).

### Characterizing malaria transmission from an asymptomatic infected individual to mosquitoes

Symptomatic *P. falciparum* is not universal, with many people in endemic areas carrying parasites in their blood without symptoms, a situation referred to as clinical immunity [27, 28]. Such individuals typically carry circulating gametocytes, and therefore can serve as reservoirs for onward transmission to mosquitoes [13, 29]. Historically, characterizing the infectiousness of asymptomatic individuals to mosquitoes has relied on mosquito feeding assays; these are costly and logistically complex, making it difficult to assess the relative importance of host factors in determining the probability of human-to-mosquito transmission. Here, we address this gap by coupling the mosquito feeding model calibrated to the mosquito feeding assay data (presented above) with a previously developed within-human parasite dynamics model that describes the temporal dynamics of asexual parasitemia and gametocytemia following blood-stage infection [18, 19] (see the Materials and Methods for details). The coupled model (referred to as the human-to-mosquito transmission model) provides a platform for linking human and mosquito factors (within-host level) with transmission probability (epidemiological level).

Fig. 4a illustrates two simulated time series of asexual parasitemia generated using the humanto-mosquito transmission model: one with parasitemia fluctuating around approximately 10^4^ parasites/mL blood—a level commonly observed in asymptomatic infections in low transmission settings [5, 30, 31], and the other with parasitemia fluctuating around approximately 10^6^ parasites/mL—a level typically observed in asymptomatic infections in high transmission settings [5, 13, 27, 31]. Details of the simulation procedure are provided in Materials and Methods. Note that the simulated time series of asexual parasitemia includes an initial exponential growth phase, consistent with observations from VIS trials [11], noting that this phase is rarely captured in field studies due to its short duration relative to typical sampling frames, followed by a transition to a steady state during which asexual parasitemia fluctuates around a relatively constant level, as observed in longitudinal cohort studies [32,33]. Fig. 4b shows, for the two simulated infections from Fig. 4a, how the probability of human-to-mosquito transmission changes over time following a bite. This probability is defined as the probability that a mosquito taking a blood meal at a particular time (in days, post infection of the human) develops oocysts (see the Materials and Methods for details). The transmission probability (per bite) curve for an individual with simulated high asexual parasitemia is much higher than that for an individual with simulated low asexual parasitemia.

**Figure 4:**
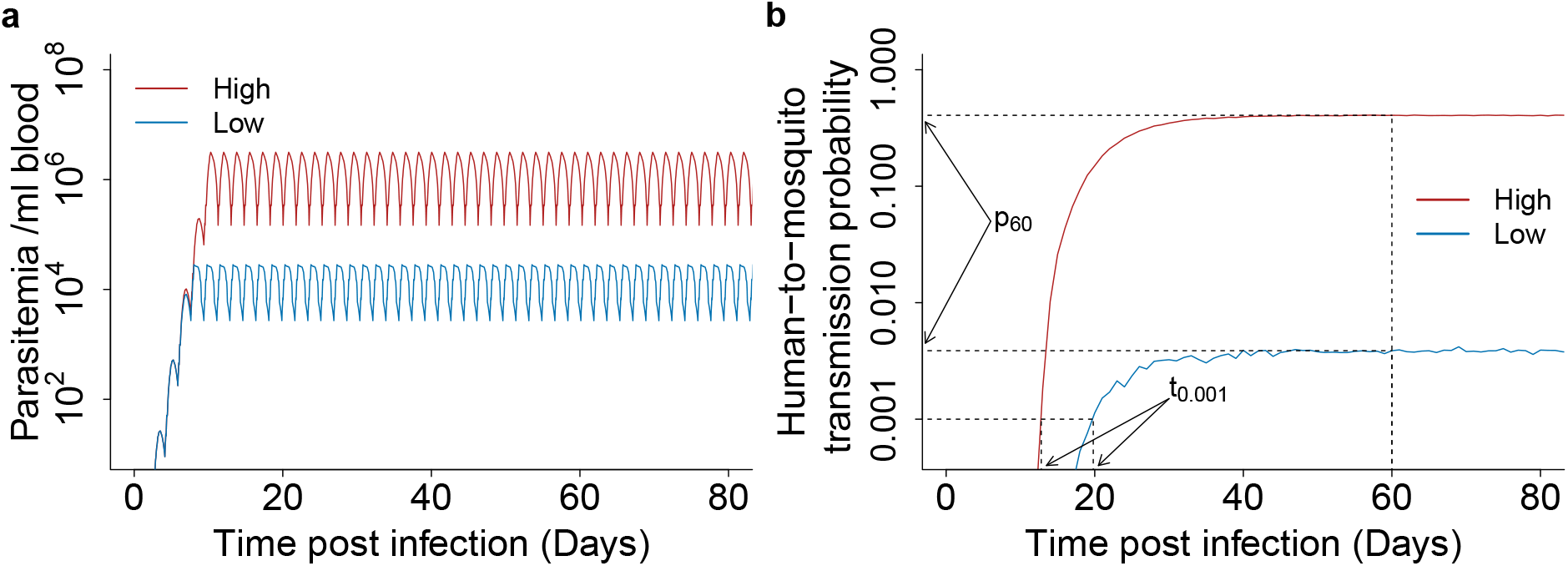
The probability of human-to-mosquito transmission of asymptomatic infections. (a) Simulated trajectories of asexual parasitemia in asymptomatic infections fluctuate around immune-regulated set-point densities of 10^4^ (low transmission settings, blue) and 10^6^ (high transmission settings, red). (b) The human-to-mosquito transmission probabilities associated with the parasitemia trajectories in (a). For each time point, we simulated the mosquito feeding model with 100,000 mosquitoes, and the transmission probability is given by the proportion of surviving mosquitoes developing oocysts (details for the simulations are provided in the section of Simulation of mosquito feeding assays and calculation of the probability of human-to-mosquito transmission in the Materials and Methods). The choice of 100,000 mosquitoes, which is substantially larger than that used in empirical mosquito feeding experiments, provides a more accurate prediction of the transmission probability to support subsequent sensitivity analysis.

The period following infection until any parasites appear in the blood is referred to as the prepatent period, while the onset of infectiousness (of the human to a mosquito) does not occur until mature gametocytes have developed (typically approximately 14 days for *P. falciparum*). While in simple models of contagiousness such as the Ross–MacDonald model, a delay between infection and infectiousness (known as the latent period) can be incorporated, the level of infectiousness is fixed throughout the infectious period [34]. In reality, and in our model (Fig. 4b), the number of gametocytes circulating increases as more are produced with rising parasitemia, until a stable level is reached. Therefore the probability (per bite) of infection increases (and may later stabilize) rather than undergoing a discrete switch in which a human changes from being non-infectious to infectious. Two quantities were defined to enable us to assess which biological parameters most strongly influence the transmission probability in the increasing and stable phases:

- The time at which the transmission probability first surpasses a specific threshold (denoted by *t*_0.001_), which characterizes the critical early (increasing) phase of the human to mosquito infection process. Specifically, we define *t*_0.001_ as the time post-infection at which the probability of a mosquito becoming infected from a single blood meal reaches 0.001 (i.e., 1 in 1000 chance of a mosquito becoming infected given a blood meal) (Fig. 4b). Using an alternative threshold (i.e., *t*_0.0005_) does not affect the results of the subsequent analysis, as will be demonstrated in the following sections whenever relevant.
- The probability of human-to-mosquito transmission at day 60 post-infection (denoted by *p*_60_), chosen as a representative time point given the prolonged duration of asymptomatic infections [29]. This quantity serves as a surrogate for the probability of human-to-mosquito transmission at steady state.

These two metrics (*t*_0.001_ and *p*_60_) effectively capture, and so summarize, how the transmission probability (and its dependence on time-since-exposure) differs between the two malaria transmission settings. They are therefore suitable metrics for use in subsequent sensitivity analyses.

### Determinants of the probability of human-to-mosquito transmission in asymptomatic infections

To investigate the relative importance of different host factors in determining the probability of human-to-mosquito transmission, a global sensitivity analysis was conducted using the partial rank correlation coefficient (PRCC) method (see Materials and Methods) [35]. The sensitivity analysis quantifies the partial rank correlations between different host factors (i.e., model parameters that capture the associated biological processes) and *t*_0.001_ or *p*_60_. Four independent PRCC analyses were performed: one using the within-human model that generates the parasitemia profile under low transmission settings (i.e., the blue curve in Fig. 4a) coupled with the posterior estimates of *ρ* and *p*_*f*_ from the Australian VIS data (which we use to approximate a low transmission setting); and three using the within-human model that generates the parasitemia profile under high transmission settings (i.e., the red curve in Fig. 4a) coupled with the posterior estimates of *ρ* and *p*_*f*_ from Burkina Faso, Cameroon, and Mali, respectively. Note that PRCC values closer to 1 or −1 indicate a stronger positive or negative correlation, respectively, whereas values closer to 0 indicate a weaker correlation. The factors with an absolute PRCC value greater than 0.5 are considered influential in the subsequent analysis.

For *t*_0.001_, Fig. 5 shows that, in the low transmission setting, it is predominantly influenced by two biological factors: the sexual conversion rate *f* and the gametocyte maturation rate *m*. In the high-transmission setting, the asexual parasite replication number *r*_*p*_ and death rate *δ*_*p*_ become the two most influential parameters, while *m* and *f* remain influential but rank lower. The result remains valid for other choices of the threshold used to define *t*_0.001_; for example, choosing *t*_0.0005_ rather than *t*_0.001_ for the PRCC analysis does not alter the findings (see S4 Fig). The marginal relationships for all parameters examined in the PRCC analysis are provided in S5 Fig–S8 Fig to provide an overview of how the samples are related and distributed. The sexual conversion rate *f* and gametocyte maturation rate *m* are not readily amenable to direct intervention, as they are largely governed by intrinsic biological processes, although sexual conversion may also be influenced by host immunity [36, 37]. The parasite replication number *r*_*p*_ and the death rate *δ*_*p*_ are the key parameters determining the multiplication of parasites per asexual replication cycle, commonly referred to as the parasite multiplication rate (PMR). Since PMR can vary significantly across individuals due to biological heterogeneity within human hosts [38], substantial variation in *t*_0.001_ is expected, even though this is challenging to observe in field studies. Mosquito-related factors are generally less influential in determining *t*_0.001_, including the two parameters we estimated earlier: the male gamete-to-gametocyte ratio *ρ* and the probability of gamete fertilization *p*_*f*_.

**Figure 5:**
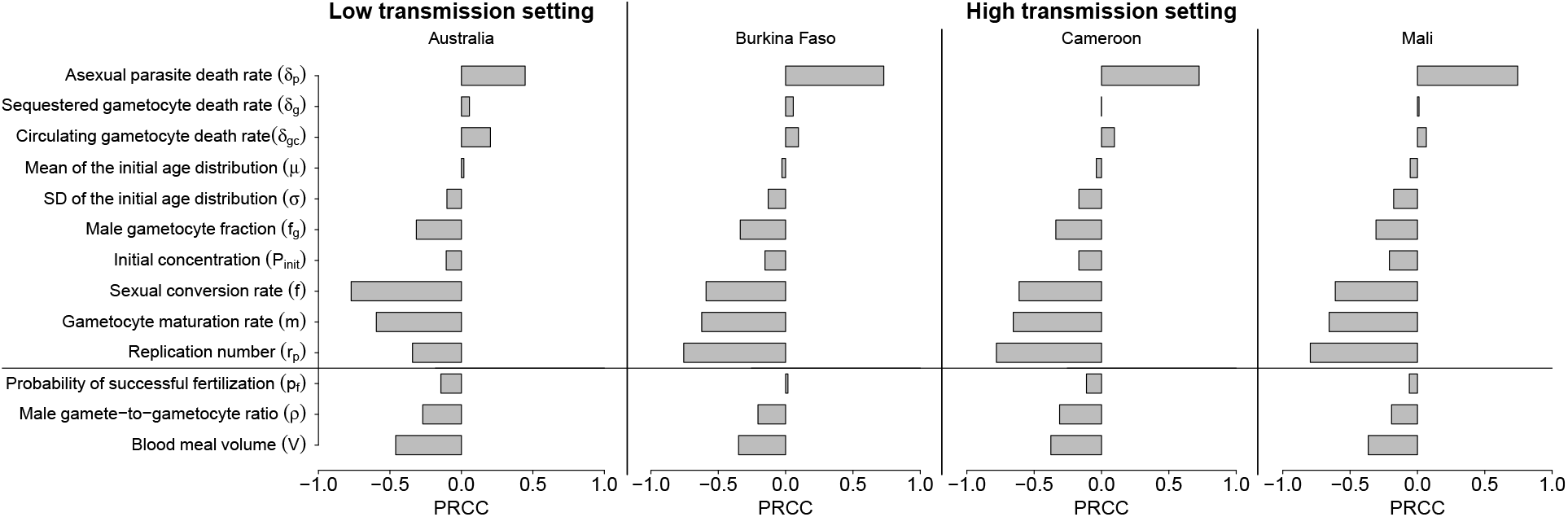
PRCC analysis of model parameters for *t*_0.001_. The parameters include those representing host factors in humans as well as those associated with mosquitoes. The human parameters were generated using Latin Hypercube Sampling (LHS), while for the mosquito parameters, *V* was sampled from a uniform distribution and *ρ* and *p*_*f*_ were jointly sampled from the posterior distributions estimated by fitting the model to the corresponding datasets. The posterior distribution estimated using the Australian dataset was used for the low transmission setting, whereas those estimated using the Burkina Faso, Cameroon, and Mali datasets were used for the high transmission setting. Details are provided in the Materials and Methods. PRCC values closer to 1 or −1 indicate stronger positive or negative correlations, respectively, whereas values closer to 0 indicate weaker correlations.

Fig. 6 shows that, in both the low and high transmission settings, *p*_60_ is predominantly influenced by four factors: three human-related parameters, namely the sexual conversion rate *f*, the fraction of male gametocytes *f*_*g*_, and the death rate of circulating gametocytes *δ*_*gc*_, and one mosquito-related parameter, namely the blood meal volume *V*. The three leading human-related factors collectively determine the availability of male gametocytes in the human bloodstream, while the mosquito blood meal volume subsequently determines the number of female and male gametocytes ingested by mosquitoes. Note that the probability of successful fertilization *p*_*f*_ and the male gamete-to-gametocyte ratio *ρ* also become influential for Cameroon. The marginal relationships for all parameters examined in the PRCC analysis are provided in S9 Fig– S12 Fig. Notably, except for *f*, all other influential factors are distinct from those that were dominant in the analysis of *t*_0.001_ (i.e., *r*_*p*_, *δ*_*p*_, *m* and *f*), indicating that the period *t*_0.001_ and the steady-state transmission probability *p*_60_ are governed by distinct biological mechanisms.

**Figure 6:**
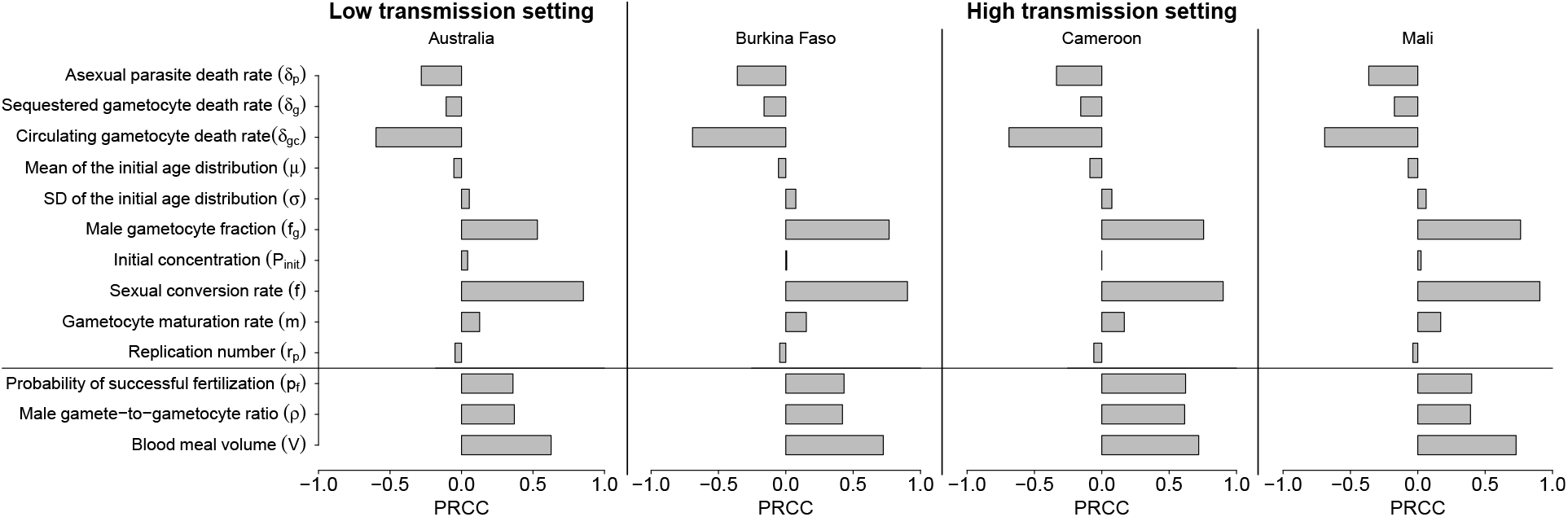
PRCC analysis of model parameters for the steady-state infectiousness *p*_60_. The parameters include those representing host factors in humans as well as those associated with mosquitoes. The human parameters were generated using Latin Hypercube Sampling (LHS), while for the mosquito parameters, *V* was sampled from a uniform distribution and *ρ* and *p*_*f*_ were jointly sampled from the posterior distributions estimated by fitting the model to the corresponding datasets. The posterior distribution estimated using the Australian dataset was used for the low transmission setting, whereas those estimated using the Burkina Faso, Cameroon, and Mali datasets were used for the high transmission setting. Details are provided in the Materials and Methods. PRCC values closer to 1 or −1 indicate stronger positive or negative correlations, respectively, whereas values closer to 0 indicate weaker correlations.

## Discussion

In this study, we have developed a multi-scale model of human-to-mosquito malaria transmission that combines parasite dynamics in both humans and mosquitoes. Through application to experimental data, we have used the model to estimate several biological parameters, thereby quantifying the effects of various human and mosquito factors on the probability of human-to-mosquito transmission. Our primary findings include: (1) the estimation of two key biological parameters, the male gamete-to-gametocyte ratio *ρ*, and the probability of successful fertilization of a female by a male gamete *p*_*f*_; (2) the characterization of the probability of parasite transmission from asymptomatic infected individuals to mosquitoes; and (3) the identification of the relative importance of different host factors in determining the probability of human-to-mosquito transmission. Our study improves the quantitative understanding of the biological determinants of malaria parasite transmission from human hosts to mosquitoes and provides a data-analytic framework that integrates mathematical models, human challenge data and statistical inference to study malaria infection and transmission.

The MAP estimate of the male gamete-to-gametocyte ratio *ρ* was approximately three- to fourfold higher in the dataset from Collins et al. using *Anopheles stephensi* than in the three African datasets from Bradley et al. using *Anopheles coluzzii, Anopheles gambiae* s.s., or their hybrid forms. The higher estimate of *ρ* may partly reflect differences in human-derived transmission-reducing immunity. Malaria-naive participants in Collins et al. were unlikely to possess naturally acquired antibodies against sexual-stage antigens, whereas such antibodies may be present in individuals in endemic settings and may reduce the number of functional male gametes available for fertilization [39]. Differences in parasite–vector compatibility may also contribute, as the efficiency of *P. falciparum* transmission has been shown to vary among combinations of parasite isolates and mosquito species; for example, Thai isolates infected *Anopheles stephensi* more efficiently than *Anopheles gambiae* in experimental comparisons [40]. The estimate of *ρ* can be used to further quantify the underlying processes that determine this parameter. Specifically, *ρ* is defined as the product of two biological parameters: the fraction of male gametocytes that are able to produce male gametes via replication (denoted by *p*_*g*_) and the number of fertile male gametes that are produced by each male gametocyte (denoted by *α*), i.e., *ρ* = *p*_*g*_*α*. If *α* is assumed to be three—based on previous findings that each male gametocyte undergoing meiosis can produce eight gametes [6] with approximately 40% or fewer of the gametes viable for fertilization [7]—we obtain a rough estimate for *p*_*g*_ of 0.27 (uncertainty: 0.043–0.97) for the dataset from Collins et al., and 0.06–0.09 (uncertainty: 0.005–0.510 for Burkina Faso, 0.003–0.59 for Cameroon and 0.005–0.51 for Mali) for the datasets from Bradley et al. This emphasizes the high uncertainty in the fraction of male gametocytes that are capable of differentiating into fertile gametes.

Our estimate for the probability of successful fertilization *p*_*f*_ between a viable macrogamete and a viable microgamete suggests that only a small proportion of viable gamete pairs undergo successful fertilization. In our model, *p*_*f*_ is defined as the probability of successful fertilization per pair of viable male and female gametes and may be interpreted as arising from either (or both) the chance that a male gamete encounters a female gamete, and that, upon encounter, fertilization occurs. While previous modeling studies have examined within-mosquito dynamics [20–22], the process of fertilization in these models is typically represented as a rate parameter (i.e., per viable gamete pair per day), with parameter values chosen to yield oocyst counts within biologically plausible ranges. To the best of our knowledge, no experimental or modeling studies have provided an *in vivo* estimate of the probability of successful fertilization for each pair of viable male and female gametes. Our study therefore advances the quantitative understanding of the fertilization process. However, our results (Fig. 3) show residual correlation in the joint posterior distributions for *ρ* and *p*_*f*_. Given that these two parameters are structurally identifiable provided the numbers of zygotes are measured, correlations are likely a result of only having presence/absence data on oocysts (i.e. the proportion of mosquitoes developing oocysts) available. Furthermore, given that the efficacy of novel vaccines likely depends upon the quantitative details of gametogenesis and fertilization (processes which are currently not fully resolved by our analyses, i.e., the residual correlations presented in the posterior estimates for *ρ* and *p*_*f*_), new mosquito feeding experiments that quantify the number of zygotes (or alternatively the number of oocysts, assuming all zygotes can settle in the mosquito midgut) would be important. In addition, mosquito immune responses may affect gamete development, reduce gamete viability, or inhibit successful fertilization [41]. These effects are currently represented implicitly through the parameters *ρ* and *p*_*f*_, but they can be explicitly incorporated into our existing model framework when studying problems that specifically require extensions of these model components.

In previous work, it has been shown that temperature can influence parasite development within mosquitoes, including oocyst intensity and the extrinsic incubation period from parasite ingestion to the appearance of salivary gland sporozoites, as well as the survival of infected mosquitoes [42]. In our mosquito feeding model, the parameter values governing mosquito mortality (*µ*_*M*_) and the gamma-distributed transition times from zygotes to oocysts and from oocysts to sporozoites (*α*_*ZO*_, *β*_*ZO*_, *α*_*OS*_, and *β*_*OS*_) were parameterized for 27^°^C [24]. The combined effects of temperature on parasite development and mosquito survival are complex and could alter the predicted probability of human-to-mosquito transmission. Additional temperature-specific data and model extensions incorporating temperature-dependent parasite development and mosquito survival would be required to evaluate these effects under different environmental conditions.

Previous longitudinal observations show that parasitemia in asymptomatic infections fluctuates around a relatively stable level over time [32, 33]—a pattern that can be interpreted in light of hypothesized host immune responses that act to reduce parasite multiplication rates [29,43]. We simulated time series of asexual parasitemia consistent with this behavior following an initial growth phase (Fig. 4A). Another dataset, from malaria-naïve neurosyphilis patients undergoing malaria therapy, exhibits a markedly different pattern of parasitemia [44, 45]. In contrast to the stabilized parasitemia commonly observed in field studies, these neurosyphilis patients, although asymptomatic, displayed multiple irregular spikes in parasitemia, with the first peak reaching approximately 10^8^ parasites/mL (a clinically relevant level), followed by progressively lower peaks in subsequent spikes. This parasitemia pattern has been hypothesized to result from the interaction between antigenic variant switching and the gradual development of variantspecific immunity [45]; as variant-specific immune responses develop, parasites preferentially switch to variants for which no specific immunity has been established. While the data from neurosyphilis patients highlight the complex interplay between host immunity and parasite biology in shaping parasitemia trajectories, they may not be a valid representation of the pattern of parasitemia observed in repeatedly exposed, clinically immune asymptomatic individuals in endemic areas. Modeling neurosyphilis patient data, and characterizing the (hypothetical) probability of human-to-mosquito transmission from such individuals, would require additional analyses including different parameter choices and modeling assumptions, and is therefore not within the scope of this work. In addition, the parameter values used to simulate asymptomatic infections were estimated from a single VIS study, in which participants were malaria-naïve, and therefore may not fully reflect the characteristics of asymptomatic infections in endemic settings. Incorporating additional data from endemic settings is therefore a future priority to improve the biological relevance of our work.

The sexual conversion rate *f* is an important parameter driving the kinetics of gametocytemia, thereby directly influencing transmission from humans to mosquitoes. Our analyses show that *f* is influential in determining *t*_0.001_ and *p*_60_ in asymptomatic infections, which suggests that a refined model of the sexual conversion process, particularly one incorporating the effects of parasite genetics, immunity and antimalarials [46–48], might be useful in further elucidating the determinants of transmission probability that are not captured in our current model. For example, some evidence suggests that parasite genetic polymorphisms associated with the development of artemisinin resistance may increase the sexual conversion rate and thus result in increased transmission efficiency. Furthermore, longitudinal observations have demonstrated that the ratio of circulating asexual parasites to gametocytes can vary over time within a single infected individual [49], suggesting temporal variation in the sexual conversion rate. Accounting for this temporal variation may improve model fidelity and should be explored in future work.

Our sensitivity analysis shows that, for *t*_0.001_, the dominant determinants are different between transmission settings. In the high transmission setting, *t*_0.001_ was primarily influenced by the replication number *r*_*p*_ and death rate of asexual parasites *δ*_*p*_, which determine the parasite multiplication rate. This finding supports the importance of prompt diagnosis and effective treatment with artemisinin-based combination therapies (ACTs), which rapidly reduce asexual parasite biomass [48]. Asexual blood-stage vaccines targeting merozoite invasion, such as RH5.1/MatrixM, may provide an additional approach by limiting blood-stage parasite replication [50]. In the low transmission setting, *t*_0.001_ is predominantly influenced by the sexual conversion rate and gametocyte maturation rate. These processes are intrinsic to parasite development and are not readily targeted directly. The effectiveness of control strategies in low transmission settings may need to focus on the more accessible determinants of *p*_60_. *p*_60_ is predominantly influenced by the density of circulating gametocytes available for uptake during a blood meal and by gamete fertilization efficiency in both transmission settings. These findings emphasize the importance of reducing mature gametocyte carriage and limiting their uptake by mosquitoes. Gametocytocidal drugs, including primaquine, methylene blue, and tafenoquine, may complement ACTs by reducing circulating gametocytes and onward transmission [51, 52]. In addition, transmission-blocking vaccines targeting sexual- and mosquito-stage antigens, such as Pfs48/45, Pfs230, and Pfs25, may inhibit gamete function, fertilization, or subsequent parasite development within mosquitoes [53]. In low transmission settings, asymptomatic infections often occur at submicroscopic parasite densities and may therefore escape routine detection [54]. Population-level strategies, such as mass drug administration using regimens with gametocytocidal activity, may help reduce both the undetected parasite reservoir and mature gametocyte carriage [55]. Our human-to-mosquito model, which maps human parasite dynamics to human infectiousness to mosquitoes, can be incorporated into a multi-scale epidemiological model of malaria transmission to predict the clinical and public health impact of these transmission-blocking strategies, providing valuable insights for optimizing and implementing effective malaria control strategies.

## Materials and Methods

### Methods for model fitting

This section includes: (1) mosquito feeding assay data used in model fitting; (2) a stochastic model of mosquito feeding, which captures the transmission of gametocytes from humans to mosquitoes and their subsequent development within the mosquito; and (3) methods of model fitting to estimate biological parameters.

#### Data

The data used in this modeling study include one dataset from a VIS conducted in Australia and three field datasets from Burkina Faso, Cameroon, and Mali (see Table. 1). The VIS dataset was obtained from Collins et al. [11], in which 17 healthy adult volunteers were inoculated with *P. falciparum*–infected red blood cells. Following inoculation of *P. falciparum* infected red blood cells, participants were monitored longitudinally, with quantitative PCR used to measure gametocytemia throughout the experiment. Mosquito feeding assays were performed for 11 of the 17 participants at three time points post inoculation to assess the transmission of gametocytes from humans to mosquitoes. At each time point, one single feeding assay was conducted using 30 laboratory-reared *Anopheles stephesi* mosquitoes. The data used in model fitting includes male and female gametocyte densities measured by PfMGET and Pfs25 qRT-PCR respectively and the proportion of mosquitoes developing oocysts detected by an 18S rDNA qPCR assay. The three field datasets from Burkina Faso, Cameroon, and Mali were obtained from Bradley et al. [12], in which mosquito feeding assays were conducted using blood collected from participants with naturally acquired *P. falciparum* gametocyte infections. The datasets included 64 assays from Burkina Faso, 13 from Cameroon, and 71 from Mali. In Burkina Faso, blood samples were offered to laboratory-reared mosquitoes comprising *Anopheles coluzzii, Anopheles gambiae s*.*s*., and hybrid forms, whereas in Cameroon and Mali, they were offered to laboratory-reared *Anopheles coluzzii* mosquitoes. The data used for model fitting included male and female gametocyte densities quantified using PfMGET and Pfs25 qRT-PCR, respectively, and the proportion of mosquitoes that developed oocysts, determined by microscopic examination of mosquito midguts dissected seven days after feeding.

#### Mosquito feeding model

A mathematical model was developed to describe the mosquito feeding experiments (referred to as the mosquito feeding model). The mosquito feeding model extends the within-mosquito model developed in [24] to include a probabilistic component to capture the process of gametocyte transmission from human to mosquito. The mosquito feeding model captures the stages of the lifecycle within the mosquito from gametocytes into oocysts and sporozoites, each of which are important developmental stages of the malaria parasite in the mosquito. Following [24], transition times from a pre-oocyst parasite stage (which is explicitly modeled as zygotes in our model) to oocysts and from oocysts to sporozoites are modeled, and the model does not include any outflow due to the death of parasites within the mosquito during the development process from zygotes to sporozoites. The oocyst or sporozoite are the target development stage for measurement in mosquito feeding assays [56–58]. Since the probability of human-to-mosquito transmission depends only on the presence of oocysts or sporozoites, rather than their abundance, the model tracks only the transition times from the establishment of viable oocysts to the onset of sporozoite production, without explicitly modeling sporozoite release from oocysts. Fig. 7 presents a schematic diagram of the model, describing the uptake of gametocytes by a mosquito and their development into sporozoites.

**Figure 7:**
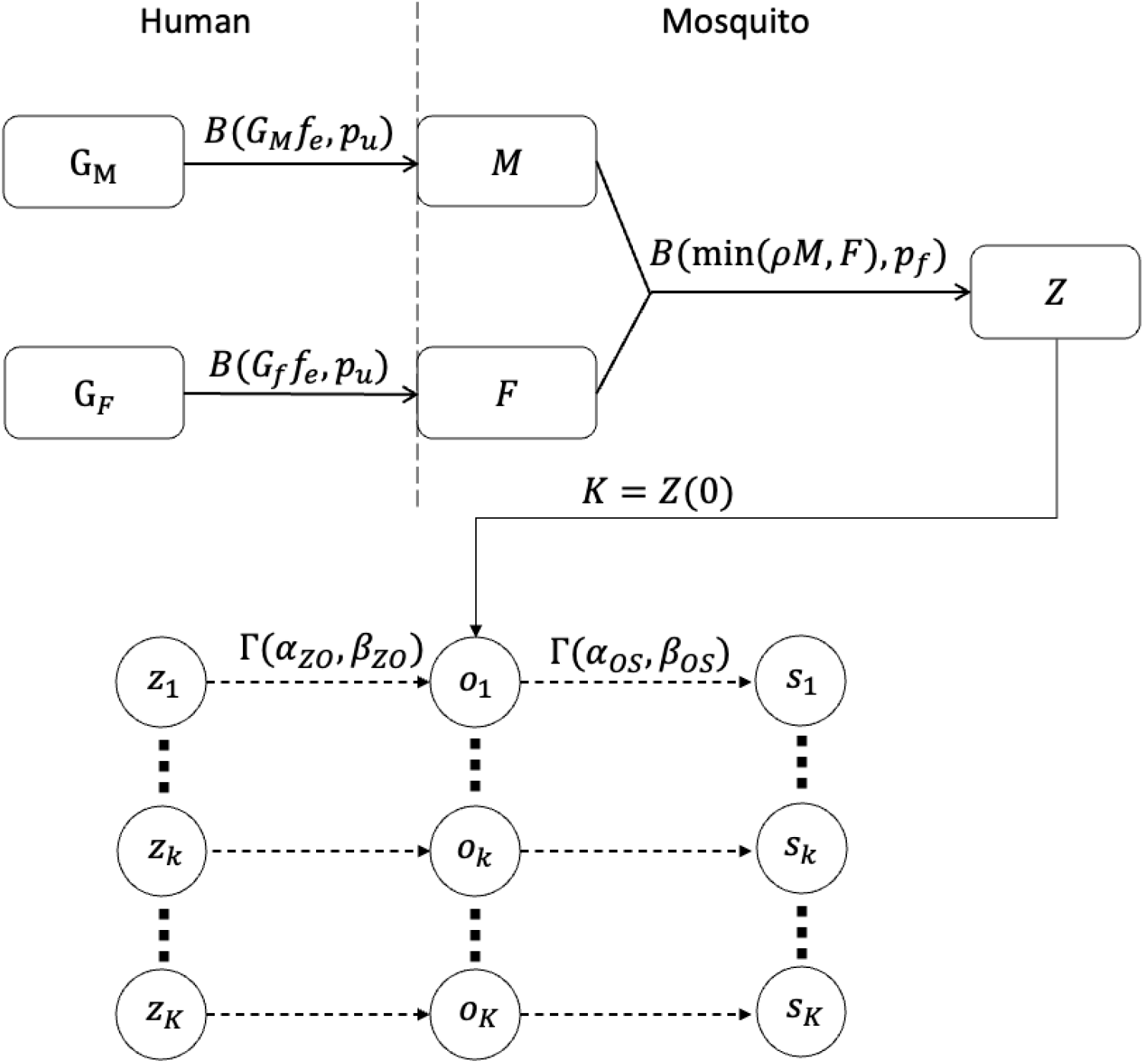
Mosquito feeding model. *M* and *F* represent the number of male and female gametocytes taken by a single mosquito during a blood meal, sampled from a binomial distribution Bin(*G* _{*M,F*}_ *f*_*e*_, *p*_*u*_), where *G*_*M*_ and *G*_*F*_ are the number of male and female gametocytes in the bitten human, *f*_*e*_ is the fraction of the total human blood volume *V*_*H*_ localized to the site of a mosquito bite (*V*_*H*_ is assumed to be 4000mL), and *p*_*u*_ is the probability of uptake by the mosquito, which is the ratio of a mosquito blood meal volume *V* to the volume of blood *f*_*e*_*V*_*H*_. *Z* represents the number of zygotes. The initial number of zygotes *Z*(*τ* = 0) is sampled from a binomial distribution Bin(min(*ρM, F*), *p*_*f*_), where *ρ* is the male gamete-to-gametocyte ratio, defined as the product of the fraction of male gametocytes that are able to produce male gametes via meiosis *p*_*g*_ and the number of male gametes that are produced by male gametocytes and viable for fertilization *α*, and *p*_*f*_ is the probability of successful fertilization for each pair of gametes. Each zygote *z*_*k*_ (*k* ∈ {1, 2, …, *K*}, *K* = *Z*(0)) develops into an oocyst *o*_*k*_ with a transition time sampled from a gamma distribution Γ(*α*_*ZO*_, *β*_*ZO*_). Each oocyst *o*_*k*_ then undergoes sporogony and starts producing sporozoites (a state modeled by *s*_*k*_) after an additional time sampled from a gamma distribution Γ(*α*_*OS*_, *β*_*OS*_).

The number of male gametocytes taken by a mosquito during a blood meal, *M*, is assumed to follow the binomial distribution:

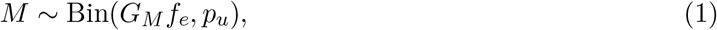

where *G*_*M*_ is the total number of male gametocytes in human body and *f*_*e*_ is the fraction of the total human blood volume *V*_*H*_ localized to the site of a mosquito bite. The total number of male gametocytes available for mosquito uptake in a blood meal is given by *G*_*M*_ *f*_*e*_, based on the assumption that circulating male gametocytes are homogeneously distributed throughout the blood. The probability of gametocyte uptake by the mosquito, *p*_*u*_, is given by *p*_*u*_ = *V/*(*f*_*e*_*V*_*H*_), which is the ratio of a mosquito blood meal volume *V* to the volume of blood *f*_*e*_*V*_*H*_ that contains *G*_*M*_ *f*_*e*_ male gametocytes (e.g., assuming a human blood volume of *V*_*H*_ = 4000mL, a value of *f*_*e*_ = 1*/*4000 implies that *f*_*e*_*V*_*H*_ = 1mL of blood is localized at the site of mosquito bite, and therefore the maximum number of male gametocyte that can be taken by mosquitoes corresponds to the number of male gametocyte in 1mL of blood). Note that *f*_*e*_ has an upper bound of 1 based on its definition and a lower bound of *V/V*_*H*_ below which the selection probability *p*_*u*_ becomes 1 and the binomial distribution model is no longer valid to capture the biological process. Given the total number of female gametocytes in human body *G*_*F*_, the number of female gametocytes taken by a mosquito during a blood meal *F* is similarly defined as follows:

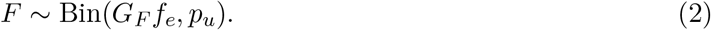

After ingestion, gametocytes develop into gametes and undergo fertilization to form zygotes which will develop into oocysts [59]. A female gametocyte only becomes a single female gamete while a male gametocyte produces a small number of male gametes via a replication process [6,7, 60]. However, not all male gametocytes ingested by mosquitoes undergo replication to produce male gametes [61], and some of the male gametes lack nuclei and are therefore not viable for fertilization [7]. Accordingly, the initial number of zygotes *Z*(*τ* = 0) is modeled by a binomial distribution:

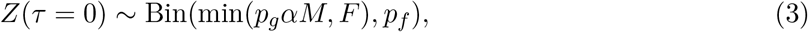

where *p*_*g*_ is the fraction of male gametocytes that are able to produce male gametes via replication, *α* is the number of male gametes that are produced by male gametocytes and viable for fertilization and *p*_*f*_ is the probability of successful fertilization. Since *p*_*g*_ and *α* are structurally non-identifiable, we define *ρ* = *p*_*g*_*α*, which represents the ratio of the number of viable male gametes to the number of male gametocytes ingested by the mosquito (i.e., the male gamete-to-gametocyte ratio). After determining the initial number of zygotes *Z*(*τ* = 0) post feeding, for each of the zygotes *z*_*k*_ (*k* ∈ {1, 2, …, *N*}, *K* = *Z*(0)), the process of the development from zygote to oocyst *O* and subsequently sporozoite *S* is modeled using an individual based model. For each zygote *z*_*k*_, transition times are sampled by randomly generating the transition times individually. The number of oocysts *O*(*τ*) at time *τ* post feeding is modeled by:

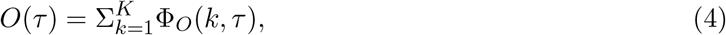

where

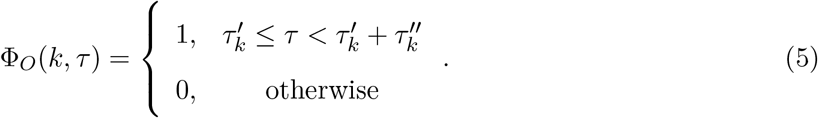

In Eq. 5, 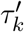 is the transition time from *Z* to *O* for the *k*th zygote, sampled from a gamma distribution Γ(*α*_*ZO*_, *β*_*ZO*_), with mean *α*_*ZO*_*/β*_*ZO*_, 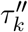 is the transition time from *O* to *S* for the *k*th zygote, sampled from a gamma distribution Γ(*α*_*OS*_, *β*_*OS*_), with mean *α*_*OS*_*/β*_*OS*_, where *α*_*ZO*_ and *α*_*OS*_ are the shape parameters and *β*_*ZO*_ and *β*_*OS*_ are the rate parameters of the gamma distributions. Since the probability of human-to-mosquito transmission depends only on the presence of oocysts or sporozoites, rather than their abundance, an indicator function was used to represent the infected status of the mosquito:

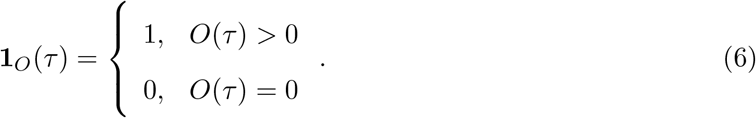

The parameter values of gamma distributions Γ(*α*_*ZO*_, *β*_*ZO*_) and Γ(*α*_*OS*_, *β*_*OS*_), which determine the times of transition between different states, are provided by [24]. Note that the mosquito-to-human transmission probability does depend on the abundance of sporozoites [62] and so future work that considered the entire transmission process would require a (straightforward) extension to this aspect of the within-mosquito model.

#### Simulation of mosquito feeding assays and calculation of the probability of human-to-mosquito transmission

This section describes the procedure for simulating a direct feeding assay in which *N*_*M*_ mosquitoes are brought into contact with an infected human whose peripheral blood contains *G*_*M*_ male gametocytes and *G*_*F*_ female gametocytes. Since the duration of mosquito feeding in the experiments is generally less than a few hours, it is reasonable to assume that gametocyte concentration in the human bloodstream does not vary during the feeding period.

The numbers of male and female gametocytes ingested by each of the *N*_*M*_ mosquitoes were simulated using Eqs. 1 and 2. For each mosquito, Eq. 3 was used to calculate the initial number of zygotes *Z*(0). The mosquito is considered to be infected if it has at least one zygote *Z*(0) ≥ 1. Eqs. 4–6 were used to simulate the times when oocysts or sporozoites are first presented in each mosquito. The mosquito is infected at time *τ* if it has developed oocysts, i.e., **1**_*O*_(*τ*) = 1.

Mosquitoes may die during the mosquito feeding assay. To account for the effect of mosquito infection status on mosquito death, a Gompertz function of time *τ* was proposed to model the mosquito death rate *µ*_*M*_. The parameters governing the Gompertz function were estimated using experimental data provided in [24], collected at 27^°^C (details about the function and data fitting are provided in the S1 Appendix).

The time of death 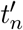 for the *n*th mosquito is determined by solving

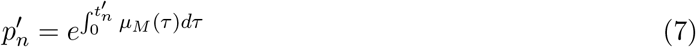

For 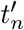, where 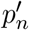 is a random number sampled from a uniform distribution *U* [0, 1]. After computing the death time 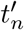 for each of the *N*_*M*_ mosquitoes, whether the *n*th mosquito survives for a duration *τ* post feeding is determined by:

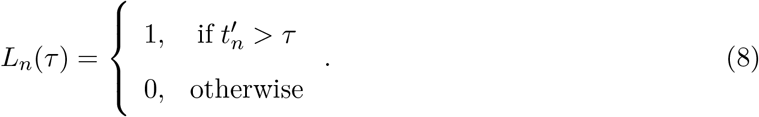

With the mortality status of each of the mosquitoes determined, the proportion of surviving mosquitoes with developed oocysts (*P*_*e*_) is given by:

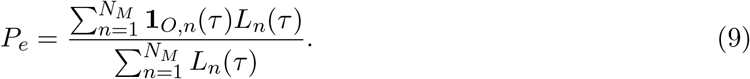

*P*_*e*_ will be fitted to the mosquito feeding assay data in two published studies [11, 12] to estimate several biological parameters (i.e., the male gamete-to-gametocyte ratio *ρ* and the probability of successful fertilization *p*_*f*_). During model fitting, *τ* was set to match the oocyst detection time in each study, with *τ* = 8.5 days for the dataset from Collins et al. [11] and *τ* = 7 days for the dataset from Bradley et al. [12]. Note that *ρ* and *p*_*f*_ are structurally identifiable provided the numbers of male and female gametocytes (*M* and *F*) and the number of zygotes (*Z*(*τ* = 0)) are available.

#### Parameters of the mosquito feeding model

All parameters of the mosquito feeding model are listed in Table 2. The value and PRCC range for the mosquito blood meal volume (*V*) were chosen from previous studies (*Anopheles stephensi*: [63]; *Anopheles coluzzii* and *Anopheles gambiae*: [64]). The value of the fraction of total human blood volume localized to the site of a mosquito bite (*f*_*e*_) is set to 0.00025, which corresponds to 1mL localized to the site of mosquito bite. Note that the value of *f*_*e*_ should be much larger than *V/V*_*H*_ (i.e., the sampling pool of the binomial distribution is equal to the blood volume *V* if *f*_*e*_ = *V/V*_*H*_) to retain stochasticity in the sampling process while also be much smaller than 1 (i.e., the sampling pool of the binomial distribution is the human blood volume *V*_*H*_) as a mosquito blood meal is a localized event and cannot capture any gametocyte circulating in the human blood stream with equal probability. We assume the mosquito bite volume *V* in mosquito feeding assay is 3.4*µ*L [63,64] and the human blood volume *V*_*H*_ is 4000mL. The expected value for the number of gametocytes taken in the blood meal by mosquitoes is independent of *f*_*e*_ and the variance only very weakly dependent on *f*_*e*_. Hence our model is highly insensitive to the choice of *f*_*e*_ (see S16 Fig). The values of the parameters governing the gamma distributions of the transition time between *Z* to *O* and *O* to *S* (i.e., *α*_*ZO*_, *β*_*ZO*_, *α*_*OS*_ and *β*_*OS*_) are chosen from [24], such that the mean transition time for zygotes to oocysts is 3.4 days and the mean transition time from oocysts to sporozoites is 9.2 days. Estimates for the male gamete-to-gametocyte ratio *ρ* and the probability of successful fertilization for each pair of gametes *p*_*f*_ are not available in the literature, and are key parameters that we estimate through our Bayesian inference procedure. The death rates of mosquitoes (*µ*_*M*_) is estimated by fitting a Gompertz function to mosquito survival data (see S1 Appendix).

**Table 2:** Parameters for the mosquito feeding model.

| Parameter | Description | Unit | Value | PRCC range | Reference |
| --- | --- | --- | --- | --- | --- |
| $f_e$ | Fraction of total human blood volume localized to the site of a mosquito bite | (unitless) | 0.00025 | fixed | See the main text |
| $V$ | Blood meal volume taken by mosquito | $\mu\text{L}$ | 3.4 | 1.3–5.4 | [63, 64] |
| $\rho$ | male gamete-to-gametocyte ratio | (unitless) | 0.8 | Sampled from joint posterior distribution | Estimated |
| $p_f$ | Probability of successful fertilization for each pair of viable gametes | (unitless) | 0.03 | Sampled from joint posterior distribution | Estimated |
| $\alpha_{ZO}$ | Shape parameter of gamma distribution for transition from $Z$ to $O$ | (unitless) | 13.8 | fixed | [24] |
| $\beta_{ZO}$ | Rate parameter of gamma distribution for transition from $Z$ to $O$ | (unitless) | 3.9 | fixed | [24] |
| $\alpha_{OS}$ | Shape parameter of gamma distribution for transition from $O$ to $S$ | (unitless) | 19.3 | fixed | [24] |
| $\beta_{OS}$ | Rate parameter of gamma distribution for transition from $O$ to $S$ | (unitless) | 2.1 | fixed | [24] |
| $\tau$ | Time for detecting oocysts post feeding | Day | 7 or 8.5 | fixed | [11, 12] |
| $\mu_M$ | The death rate of mosquitoes | Function of time | | | See S1 Appendix |

#### Model fitting

To estimate the male gamete-to-gametocyte ratio *ρ* and the probability of successful fertilization *p*_*f*_, the mosquito feeding model was fitted to the mosquito feeding assay data from previously published studies [11, 64] using an approximate Bayesian computation sequential Monte Carlo (ABC-SMC) algorithm [65]. The data used in the model fitting includes the male and female gametocytemia (*G*_*M*_ *f*_*e*_ and *G*_*F*_ *f*_*e*_ in Eqs. 1 and 2) and the proportion of mosquitoes developing oocysts (*P*_*e*_ in Eq. 9). Data from each country were pooled for model fitting (rather than evaluated using hierarchical modeling), because the parameters to be estimated are not influenced by between-volunteer variance.

In the ABC-SMC method, the posterior distribution is approximated by comparing the distance between simulated and observed data, with parameter samples initially drawn from the prior distribution and then from a sequence of intermediate distributions, progressively refined using sequential Monte Carlo, and with tolerance gradually decreasing over generations [65]. The prior distributions of *ρ* and *p*_*f*_ must be defined first. To inform the prior distribution for *ρ*, we consider that *ρ* = *p*_*g*_*α*, where *p*_*g*_ represents the fraction of male gametocytes undergoing replication to produce male gametes and *α* is the mean number of viable male gametes produced per male gametocytes undergoing replication. There is substantial uncertainty surrounding the value of *p*_*g*_, with experimental studies reporting values spanning the full range from 0 to 1 [61,66]. For *α*, it is known that each male gametocyte undergoing replication gives rise to 8 male gametes [6], but not all of these are viable, as only gametes containing a nucleus are capable of fertilization [7]. Given that at least one nucleus is present per gametocyte, the plausible biological range for *α* is between 1 and 8. Accordingly, the prior distribution for *ρ* is given by *U* [0, 1] *× U* [1, 8] (see S2 Appendix). For *p*_*f*_, where limited empirical information is available, a uniform prior distribution *U* [0, 1] was chosen, allowing the data to inform the posterior distribution.

In the first generation of the ABC-SMC algorithm, after specifying the prior distributions, the parameters *ρ* and *p*_*f*_ are sampled independently from prior distributions. In subsequent generations, *ρ* and *p*_*f*_ are sampled from the intermediate posterior distributions. For each parameter sample (*ρ, p*_*f*_), simulated data 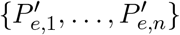 are generated by using Eqs. 1–9, where *n* is the number of data points (see Table 1). The Euclidean distance *d* between the simulated data 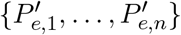 and observed data {*P*_*e*,1_, …, *P*_*e,n*_} is given by:

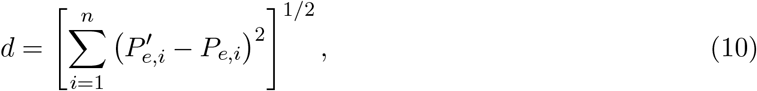

which is a common choice of distance metric (see [67] for details). The sampling process is repeated until 5000 samples are accepted in each generation. The algorithm terminates when the acceptance rate falls below 0.5%, which serves as the stopping rule [68]. Full details of the ABC-SMC implementation, including the choice of tolerance, resampling and weighting, and pseudo-code, are provided in S2 Appendix. Fitting results include the joint posterior distribution of *ρ* and *p*_*f*_ and the relationship between the proportion of mosquitoes developing oocysts and male and female gametocytes generated by 5000 model simulations based on 5000 samples from the posterior parameter distribution (Fig. 2).

#### A model of within-human parasite dynamics

Here we introduce a new model of within-human parasite dynamics that is integrated with the mosquito feeding model to form the multi-scale human-to-mosquito transmission model. The within-human parasite dynamics model was developed by extending a previously developed gametocyte dynamics model [18,19] to capture the development of male and female gametocytes. A schematic diagram of the new within-human parasite dynamics model is shown in Fig. 8. The model contains three major parasite populations: asexual parasites *P* (*a, t*), sexually committed parasites *P*_*G*_(*a, t*) and gametocytes (including immature stages *G*_1_(*t*)–*G*_4_(*t*) and mature male *G*_*M*_ (*t*) and female *G*_*F*_ (*t*) populations), where *a* represents parasite age and *t* represents time since the onset of blood stage infection. Asexual parasites can invade red blood cells (RBCs) and initiate an asexual life cycle lasting *a*_*L*_ hours, during which they undergo maturation and replication. The parasitized RBCs (pRBC) may die at any time governed by a rate parameter *δ*_*p*_. If they reach the end of the asexual life cycle, rupture occurs, releasing merozoites which invade uninfected RBCs and initiate a new replication cycle. During the asexual life cycle, a small fraction of the parasites (at age *a*_*s*_) differentiate into sexually committed parasites and eventually develop into male and female gametocytes.

**Figure 8:**
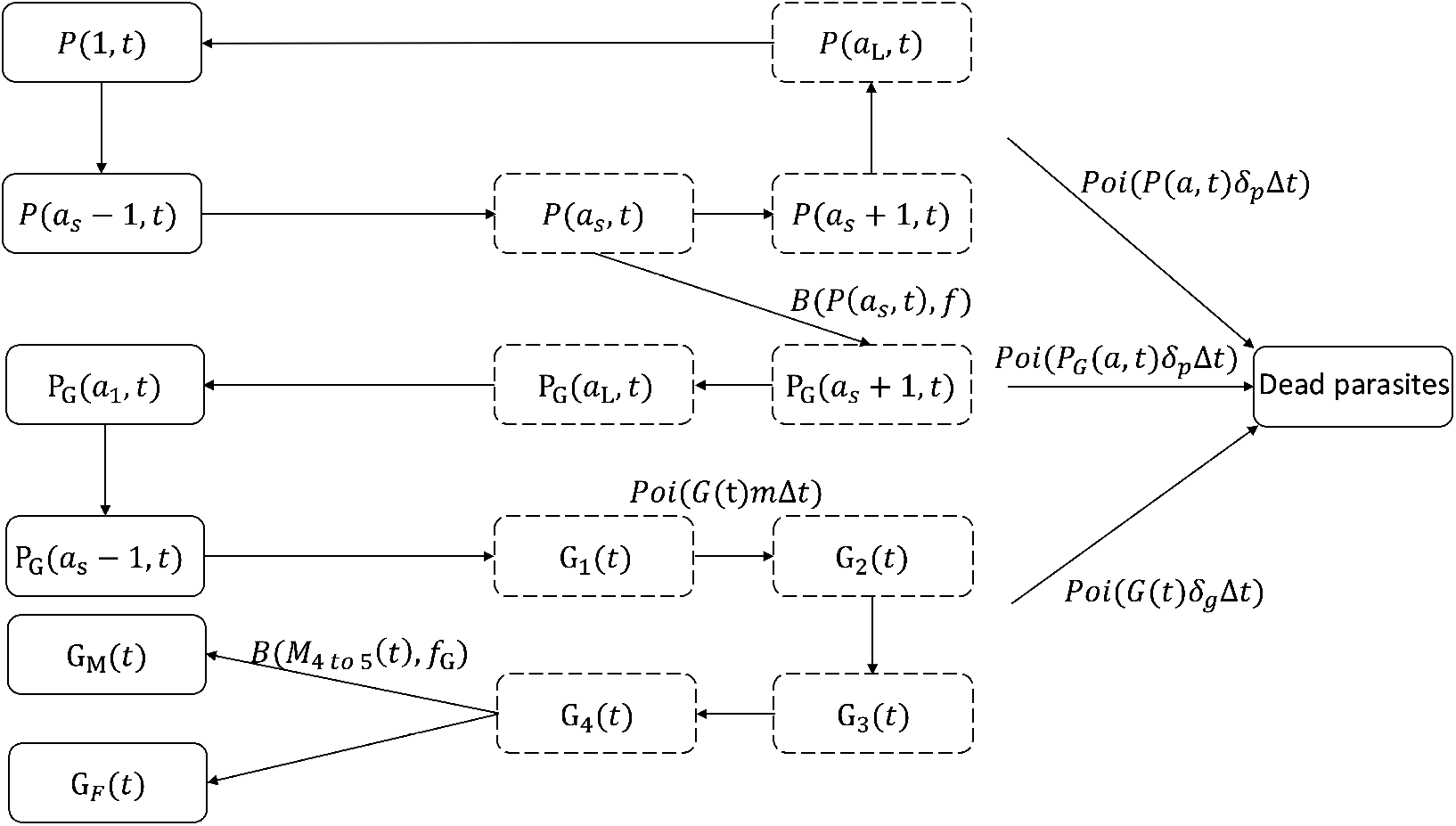
A schematics diagram of the new within-human parasite dynamics model. The model contains three major parasite populations: *P* (asexual parasites), *P*_*G*_ (sexually committed parasites) and *G* (gametocytes). *P* and *P*_*G*_ are functions of parasitemia with age *a* at time *t*. The loop *P* (*a, t*) describes the synchronous waves of asexual replication and maturation in *a*_*L*_ hours. A small fraction *f* of asexual parasites enter the sexual development cycle when sexual commitment occurs at age *a*_*s*_, developing into sequestered gametocytes *G*_1_(*t*)–*G*_4_(*t*) and eventually becoming circulating mature male (*G*_*M*_) and female (*G*_*F*_) gametocytes, with probabilities *f*_*G*_ and 1 − *f*_*G*_ respectively. Parasites may die at any time point during the developmental process, following a Poisson process with a probability per unit time (i.e., per time step Δ*t* of one hour) determined by *P* (*a, t*)*δ*_*p*_Δ*t* (for asexual parasites), *P*_*G*_(*a, t*)*δ*_*p*_Δ*t* (for sexually committed parasites) and *G*(*t*)*δ*_*g*_Δ*t* (for gametocytes), where *δ*_*p*_ is the natural death rate of parasites and *δ*_*g*_ is the natural death rate of gametocytes. Compartments with a dashed boundary represent those for which parasites of a particular age are sequestered to tissues and thus not measurable by sampling peripheral blood, whereas parasitemia refers to the parasite density in compartments with solid boundaries.

The within-human parasite dynamics are described by a discrete-time stochastic model with a time step of Δ*t* = 1 hour. A tau-leaping algorithm [69] was used to simulate this model. In detail, the total number of asexual parasites at the time of inoculation *P* (*a*, 0) was first calculated by multiplying the initial density parameter *P*_*init*_ (see Table 3) by an assumed blood volume *V*_*H*_ of 4000mL. The total number of asexual parasites *P* (*a*, 0) was then distributed across the *a*_*L*_ age bins by generating *P* (*a*, 0) random samples from a truncated normal distribution 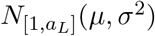 and rounding the samples to the nearest integers. Subsequently, the number of asexual parasites changes according to the following equation:

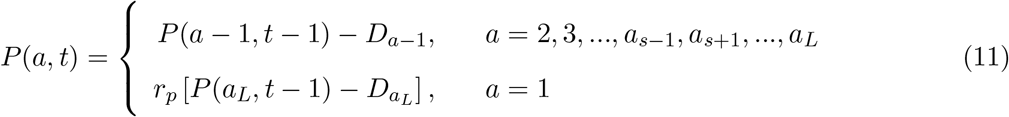

where *a*_*L*_ is the duration (in hours) of each asexual replication cycle, *a*_*s*_ is the age at which sexual commitment occurs, *r*_*p*_ is the parasite replication number indicating the average number of newly infected RBCs attributable to the rupture of a single infected RBC. Note that the parasite replication number *r*_*p*_ is different from the so-called parasite multiplication rate, the latter of which is a ‘net replication rate’ quantified by the increase in parasite numbers due to replication (*r*_*p*_) and the decrease due to parasite death or sexual commitment. Parasite multiplication rate is calculated by 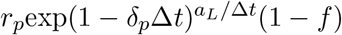, where the term 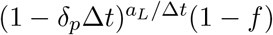 gives the fraction of surviving asexual parasites after death and sexual conversion per replication cycle (see S3 Appendix for the details), *δ*_*p*_ is the rate of natural death of asexual parasites and *f* is the probability of sexual commitment for each parasite. The number of asexual parasites that die *D*_*a*_ during each time step (Δ*t* = 1h) is sampled from a Poisson distribution Poi(*P* (*a*− 1, *t*− 1)*δ*_*p*_Δ*t*).

**Table 3:** Parameters for the within-human parasite dynamics model.

| Parameter | Description | Unit | Value | PRCC range | Reference |
| --- | --- | --- | --- | --- | --- |
| $P_{init}$ | Initial concentration | parasite/mL | 0.08 | 0.05–0.15 | [18] |
| $\mu$ | Mean of the initial age distribution | h | 7 | 7–12 | [18] |
| $\sigma$ | SD of the initial age distribution | h | 8 | 4–12 | [70] |
| $r_p$ | Replication number | (unitless) | 71 | 20–100 | [18] |
| $f$ | Fraction of parasites entering sexual development | (unitless) | 0.006 | 0.003–0.1 | [18, 71, 72] |
| $\delta_p$ | Death rate of asexual and sexual parasites | $h^{-1}$ | 0.03 | 0.01–0.04 | [18] |
| $\delta_g$ | Death rate of sequestered gametocytes | $h^{-1}$ | 0.0004 | 0.0003–0.001 | [18] |
| $\delta_{gc}$ | Death rate of circulating gametocytes | $h^{-1}$ | 0.007 | 0.003–0.01 | [18] |
| $m$ | Maturation rate of gametocytes | $h^{-1}$ | 0.02 | 0.01–0.03 | [18] |
| $f_G$ | Male gametocyte fraction | (unitless) | 0.2 | 0.1–0.5 | [11, 74] |
| $a_s$ | Sequestration age of asexual parasites | h | 25 | fixed | [18] |
| $a_L$ | Length of life cycle of asexual parasites | h | 42 | fixed | [18] |

For those parasites that may undergo sexual commitment (i.e., parasites with age *a*_*s*_):

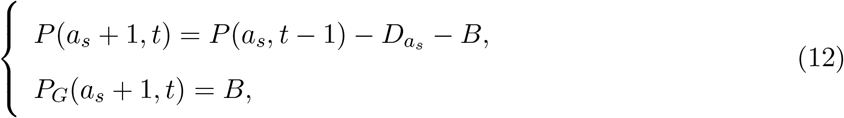

where *B* represents the number of parasites that become sexually-committed, sampled from a binomial distribution 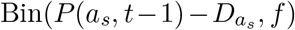 where *f* is the probability of sexual commitment for each parasite and is assumed to be fixed over the course of infection. The first line of Eq. 12 captures the evolution of the parasites remaining in the asexual life cycle, while the second line of Eq. 12 describes the number of parasites undergoing sexual commitment. Sexually committed parasites continue the rest of the replication cycle, which is modeled by:

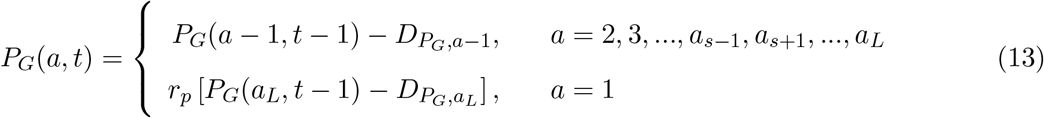

where 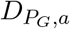 represents the number of sexually-committed parasites that die in each time step, sampled from Poi(*P*_*G*_(*a, t* − 1)*δ*_*p*_Δ*t*). The sexually committed parasites will develop into immature gametocyte stages (*G*_1_–*G*_4_) and eventually mature male gametocytes *G*_*M*_ and female gametocytes *G*_*F*_, both of which circulate in the bloodstream. The dynamics of the gametocytes *G*_1_ − *G*_4_ are modeled by:

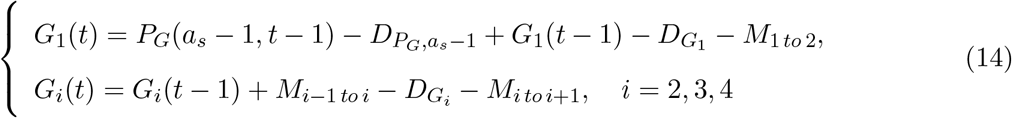

where 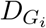 represents the number of dead stage *i* gametocytes, sampled from Poi(*G*_*i*_(*t*−1)*δ*_*g*_Δ*t*), where *δ*_*g*_ is the death rate of immature gametocytes and *M*_*i to i*+1_ represents the number of stage *i* gametocytes maturing to become stage *i* + 1 gametocytes, sampled from Poi(*G*_*i*_(*t* − 1)*m*Δ*t*) for *i* = 1, 2, 3, 4, where *m* is the rate of gametocyte maturation. Male and female gametocytes develop after stage IV [2] and their dynamics are modeled by:

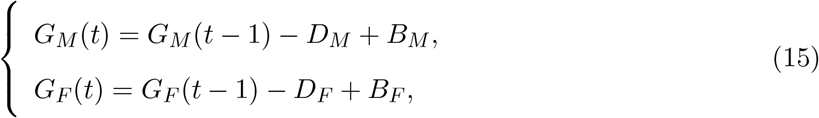

where *D*_*M*_ represents the number of dying male gametocytes during a time step, sampled from Poi(*G*_*M*_ (*t* − 1)*δ*_*gc*_), where *δ*_*gc*_ is the death rate of circulating gametocytes. Similarly, *D*_*F*_ represents the number of dying female gametocytes during a time step, sampled from Poi(*G*_*F*_ (*t* − 1)*δ*_*gc*_), *B*_*M*_ represents the number of immature gametocytes that become male gametocytes, sampled from Bin(*M*_4 *to* 5_, *f*_*G*_), where *f*_*G*_ is the probability that a sequestered *G*_4_ gametocyte develops into a male gametocyte, referred to as the male gametocyte fraction, and *B*_*F*_ represents the number of immature gametocytes that become female gametocytes, given by *M*_4 *to* 5_ − *B*_*M*_.

To simulate asymptomatic infections, the parasite replication number was adjusted to be

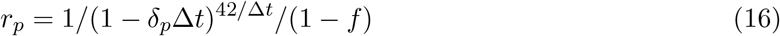

such that the PMR is equal to 1 at the specified set-point parasitemia level (i.e., 10^4^ and 10^6^), which yields cyclical asexual parasitemia around the set-point level, mimicking an active immune response.

### Methods for sensitivity analysis

To quantify the correlations between various host factors and the probability of human-to-mosquito transmission of asymptomatic infections, a partial rank correlation coefficient (PRCC) analysis was conducted. The PRCC analysis is designed to identify the correlation of two variables after removing any linear dependence on all other variables in a model with multiple parameters [35]. In detail, a total of 1000 parameter sets were generated for the sensitivity analysis. For the human parameters, Latin Hypercube Sampling (LHS) was used by sampling from joint uniform distributions, with each parameter varied within the range shown in Table 3. For the mosquito parameters, the blood meal volume *V* was sampled from a uniform distribution within its specified range (see Table 2) and *ρ* and *p*_*f*_ were randomly sampled from the 5000 joint posterior samples obtained from the Bayesian analysis. For each parameter set, the within-human parasite dynamics model was simulated once to generate the time series of gametocytemia of asymptomatic infections. At each time point, the male and female gametocyte density were used as inputs in the mosquito feeding model to compute the probability of human-to-mosquito transmission defined as the proportion of surviving mosquitoes with developed oocysts (*P*_*e*_). After obtaining the time series of the transmission probability, *t*_0.001_ and *p*_60_ were calculated. The PRCC analysis was performed separately using *ρ* and *p*_*f*_ sampled from each of the joint posterior distributions reported in Table 1. The joint posterior distribution estimated from the Australian dataset was used to represent the low transmission setting, whereas those estimated from the Burkina Faso, Cameroon, and Mali datasets were used to represent the high transmission setting. The results of the analysis include the PRCC values between *t*_0.001_ and the parameters shown in Fig. 5, as well as between *p*_60_ and the same parameters shown in Fig. 6.

Using a single simulation for each parameter set may introduce variability into the PRCC results in the stochastic model. Previous studies have shown that stable PRCC results can be obtained when the number of sampled parameter sets is sufficiently large [35]. To evaluate the robustness of the primary analysis, the PRCC analysis for low transmission setting was repeated using the same 1000 parameter sets, with 100 stochastic replicates performed for each set. The resulting values of *t*_0.001_ and *p*_60_ were averaged across replicates. The results are provided in S13 Fig– S15 Fig.

### Parameters of the within-human parasite dynamics model

All parameters of the within-human parasite dynamics model are listed in Table 3. The parameter values and PRCC ranges for the within-human parasite dynamics model are primarily based on population-level estimates reported in [18], where a gametocyte dynamics model was fitted to VIS data [11]. The PRCC ranges are selected to cover the 95% HDPI of the parameter estimates. The SD of the initial age distribution (*σ*) is instead taken from [70], as volunteers in the VIS were inoculated with predominantly early ring-stage parasites, resulting in a tighter parasite age distribution than in natural infections. The PRCC range of the sexual conversion rate (*f*) is chosen from [71, 72]. The value and PRCC range of the male gametocyte fraction (*f*_*G*_) are chosen from [11, 73, 74].

All the parameters of the within-human parasite dynamics model are included in the partial rank correlation coefficient (PRCC) sensitivity analysis, except for the sequestration age of asexual parasites *a*_*s*_ and the length of the replication cycle of asexual parasites *a*_*L*_. For the parameters of mosquito feeding model, the blood meal volume taken by mosquito *V*, the male gamete-to-gametocyte ratio *ρ* and the probability of successful fertilization for each pair of viable gametes *p*_*f*_, are included in the PRCC sensitivity analysis (see Table 2).

## Data Availability

All data produced in the present work are contained in the manuscript

## Data availability

The data used in this study were obtained from two previously published studies [11, 12] and are publicly available from the original publications. A curated version used for model fitting is provided in our repository at https://github.com/XSSUN2/Modelling-the-transmission-of-malaria-parasites-from-a-human-host-to-mosquitoes.git.

## Code availability

All the code used for model analysis and result visualization is publicly available at https://github.com/XSSUN2/Modelling-the-transmission-of-malaria-parasites-from-a-human-host-to-mosquitoes.git. All analyses were performed in R (version 4.2.1).

## Acknowledgments

We thank the Australian Research Council (ARC) for supporting this research through a Discovery Project (DP210101920) and a Laureate Fellowship awarded to J.M.M. (FL240100126) which supports X.S., P.C. and J.M.M. We thank Medicines for Malaria Venture for funding the VIS clinical trials. J.S.M. is supported by an NHMRC Investigator Grant (2016396). Computer simulations were conducted on the Nectar Research Cloud, a collaborative Australian research platform supported by the National Collaborative Research Infrastructure Strategy (NCRIS).

## Author contributions

X.S., M.W.A.D., J.S.M., J.M.M. and P.C. contributed to conceptualization. M.W.A.D. and J.S.M. contributed to data curation. X.S. contributed to formal analysis. X.S., J.M.M. and P.C. contributed to methodology. J.M.M. and P.C. contributed to supervision. X.S. contributed to visualization. X.S., J.M.M. and P.C. contributed to writing of the original draft. X.S., M.W.A.D., J.S.M., J.M.M. and P.C. contributed to review and editing of the manuscript.

## Supplementary materials

## S1 Appendix

### Estimation of death rate of mosquitoes

Previous analyses on mosquito survival have revealed that a Gompertz function provides an excellent fit to data. Mortality rates generally increase with age for mosquitoes [24, 57, 75]. In our model, the death rate *µ* is assumed to be a Gompertz function of time *τ*. We allow for a potential difference between infected (*E* = 1) and uninfected (*E* = 0) mosquitoes:

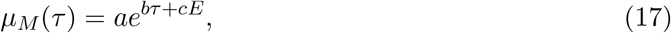

where *a, b* and *c* are non-zero constants. The mosquito survival *p*(*τ*) is then given by the following differential equation:

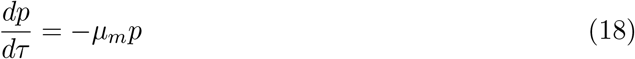

To solve this, we separate the variables and integrate both sides:

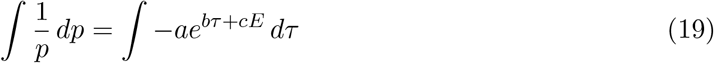

The equation integrates to:

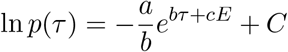

Substitute *p*(0) = 1:

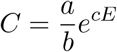

Finally, we get the solution:

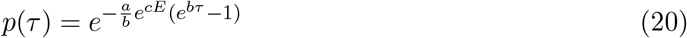

We fit this equation to mosquito survival data (at 27°C in [24]) by using nonlinear least squares estimation *nlsLM* in package *minpack*.*lm* (with default settings) in R (version 4.2.1). Results are shown in the Figure below.

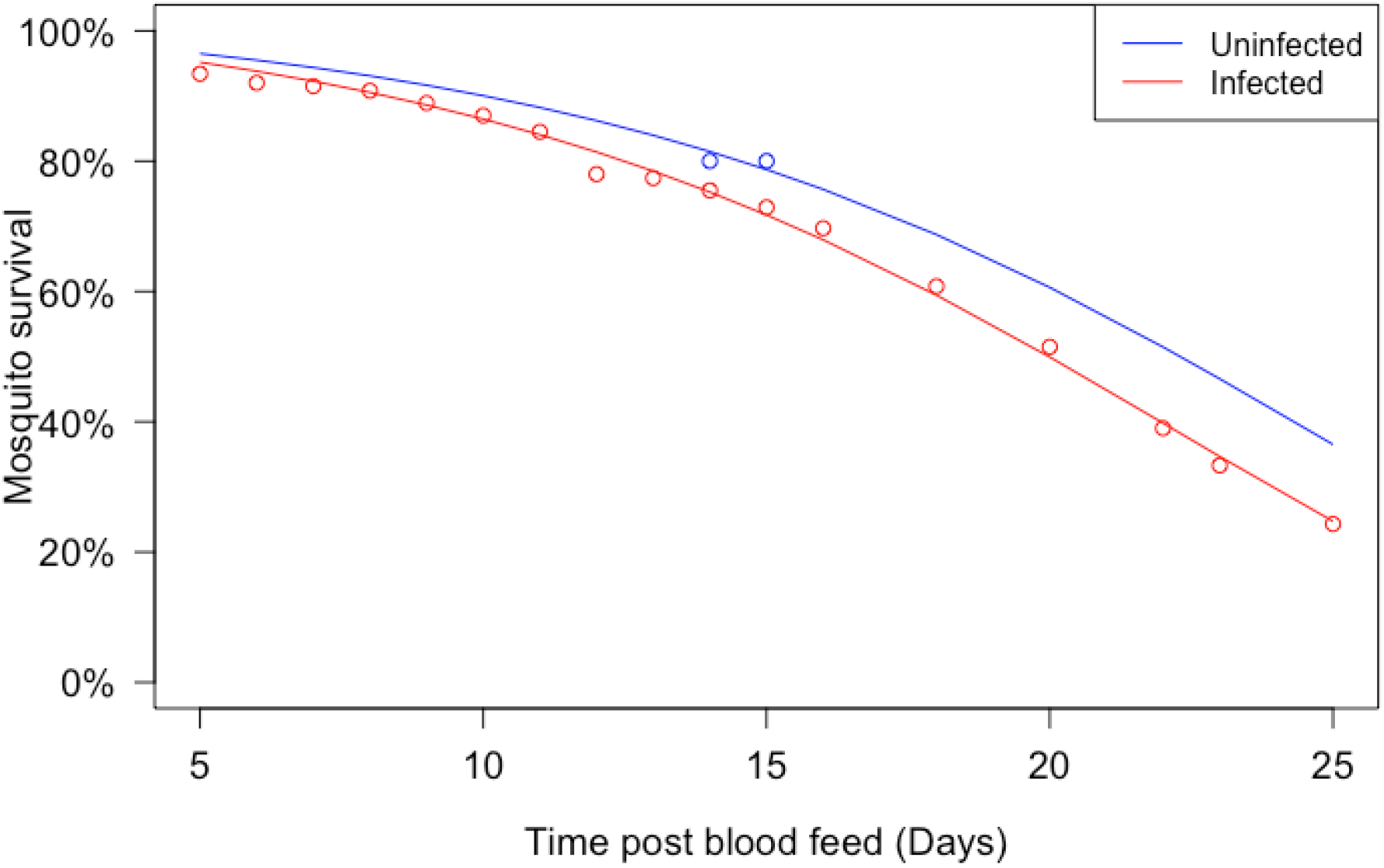

### Fitting results for the death rate of mosquitoes *µ*_*M*_

The dots represent the data for infected mosquitoes (red) and uninfected mosquitoes (blue). The curves are the fitting results for the death rate *µ*_*M*_ (infected *E* = 1, red, and uninfected *E* = 0, blue).

## S2 Appendix

### Methods for estimating biological parameters

To calculate the density of the prior distribution of *ρ, U* [0, 1] *× U* [1, 8], we employ the probability density function (PDF) transformation method. Let *X* ~ *U* [0, 1] and *Y* ~ *U* [1, 8], and define *Z* = *XY*. the individual PDFs of *X* and *Y* are:

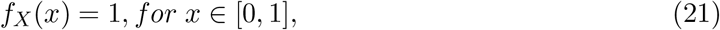

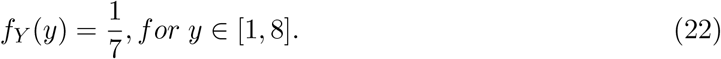

Since *X* and *Y* are independent, the joint PDF of *X* and *Y*, denoted by *f*_*X,Y*_ (*x, y*), is:

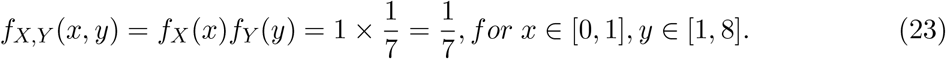

We are interested in the distribution of *Z* = *XY*. To derive *f*_*Z*_(*z*), we apply the change-of-variable technique, where the Jacobian determinant is:

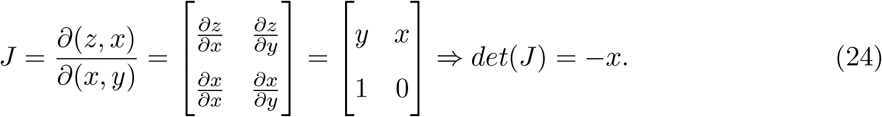

Using this transformation, the PDF of *Z* is given by:

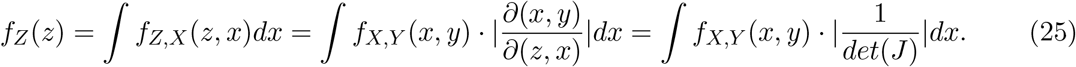

Since 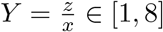, it follows that *x* ∈ [*z/*8, *min*(*z*, 1)], and the PDF of Z becomes:

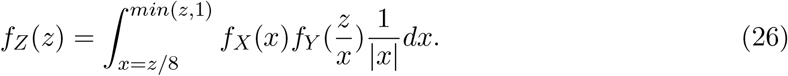

For *z* ∈ [0, 1], the PDF is:

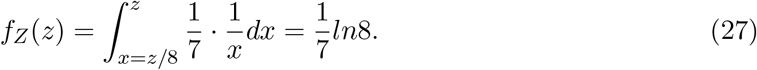

For *z* ∈ (1, 8], the PDF is:

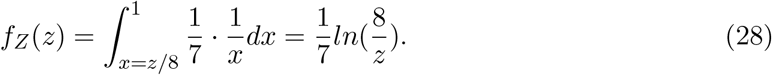

Therefore, the PDF of *Z* is:

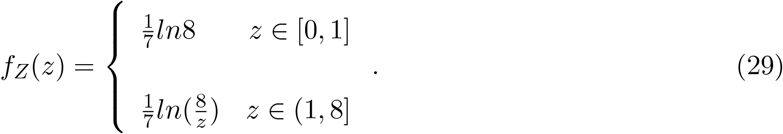

To perform the ABC-SMC, we independently sample the parameters *ρ* and *p*_*f*_ from prior distributions in the first generation (*g* = 1). For each parameter sample (*ρ, p*_*f*_), simulated data 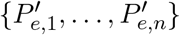 are generated based on the male and female gametocytemia. We then compute the Euclidean distance *d*, which is a common choice of distance metric (see [67] for details), between the simulated data 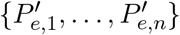 and observed data {*P*_*e*,1_, …, *P*_*e,n*_}, as follows:

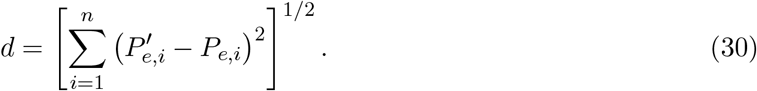

The sampled parameter pairs (*ρ, p*_*f*_) are accepted in the first generation if *d < ϵ*_1_, where 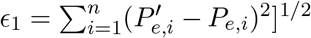 represents the ‘data-only’ distance. We repeat the sampling process until *L* = 5000 samples 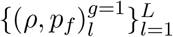 are obtained and assigned an equal normalized weight to each accepted sample 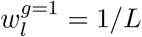. For subsequent generations, new samples (*ρ, p*_*f*_)^∗^ are drawn from the weighted empirical distribution of accepted samples from the previous generation, 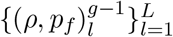, using weights 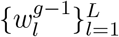. Each sample is perturbed using a multivariate normal kernel *K*[(*ρ, p*_*f*_)|(*ρ, p*_*f*_)^∗^], with mean and standard deviation set to be the sample mean and standard deviation of the accepted samples from the previous generation *g* − 1 to improve sampling efficiency [76]. As in the first generation, simulated data are generated by the model for each perturbed sample (*ρ, p*_*f*_) and the distance *d* between the simulated and observed data is calculated. (*ρ, p*_*f*_) is accepted if *d < ϵ*_*g*_, where *ϵ*_*g*_ is given by:

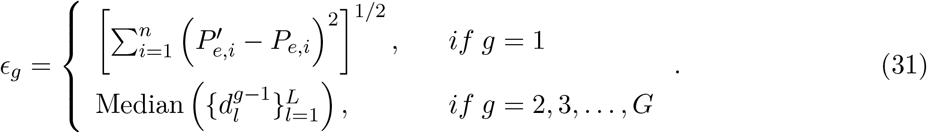

The process is repeated until *L* = 5000 samples 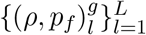 are accepted in each generation. The weights *w*^∗^ for accepted samples are calculated using:

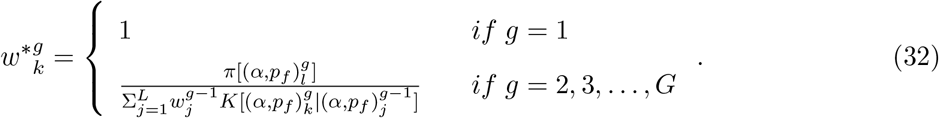

where *π* denotes the prior distributions of *ρ* and *p*_*f*_. After computing the weights, they are normalized, 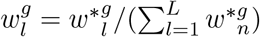, so that 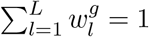. The total number of generations *G* is determined by the acceptance rate of each generation, with the algorithm terminating once the acceptance rate drop below 0.5%, which serves as the stopping rule [68]. The pseudo-code for the algorithm is provided below.

#### Algorithm 1

Pseudo-code of ABC-SMC Algorithm

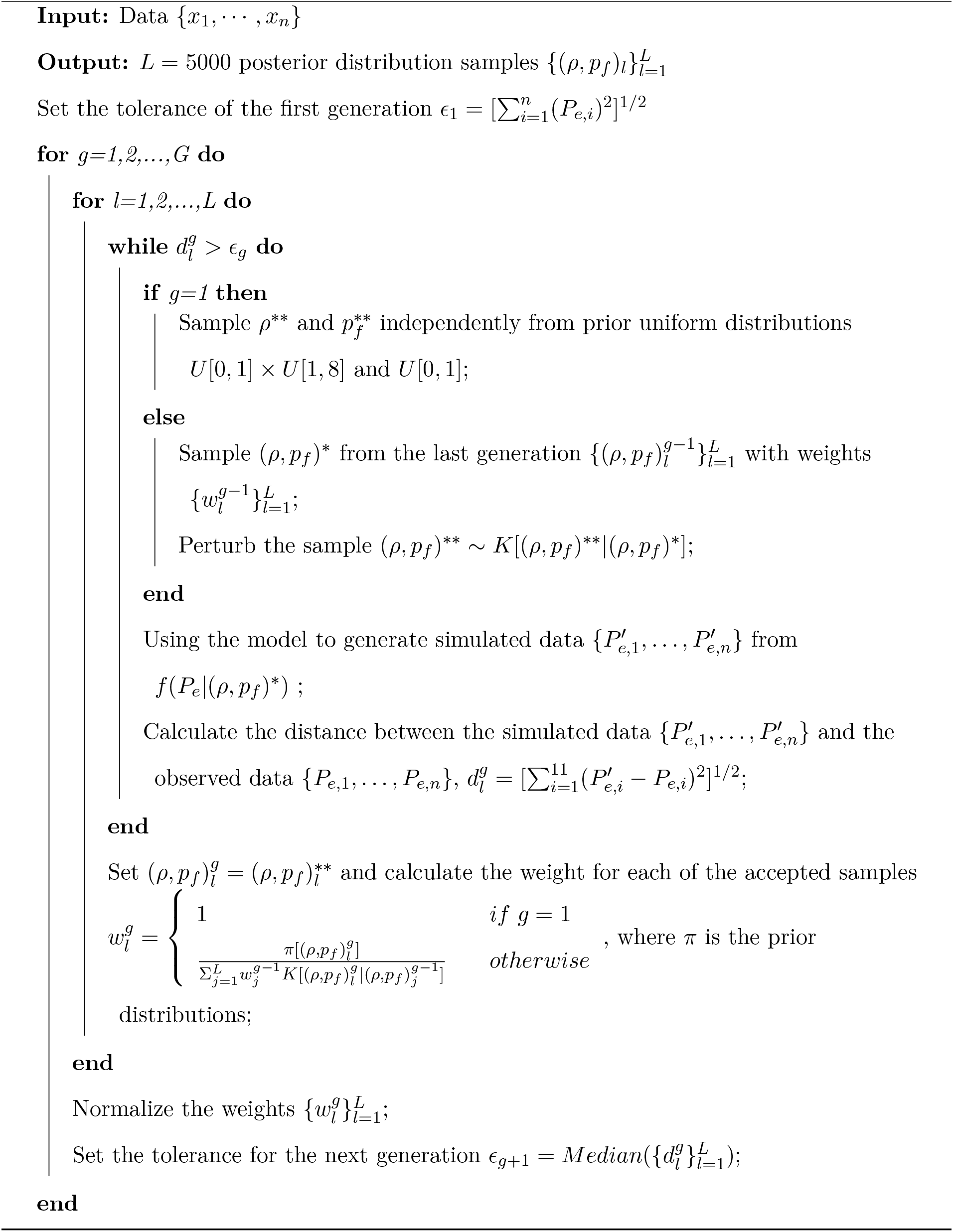

## S3 Appendix

### Method for calculating of parasite multiplication rate

Consider a death process with death rate *δ*_*p*_ with population size *P* and a single time step Δ*t* = 1 hour. Under one-step Tau-leaping, the number of deaths *D* during the step is sampled as

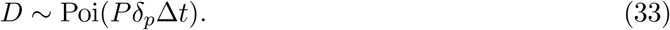

Define the (random) one-step survival fraction under Tau-leaping by

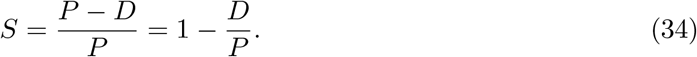

Taking expectation and using linearity of expectation gives

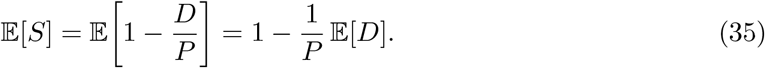

For a Poisson random variable, E[*D*] = *Pδ*_*p*_Δ*t*, hence

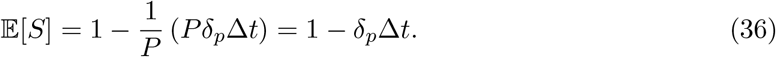

Now define a new death rate 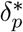 by matching the continuous-time exponential survival probability over one time step to the mean Tau-leaping survival fraction:

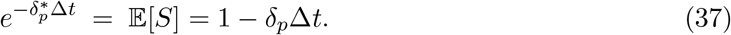

and therefore

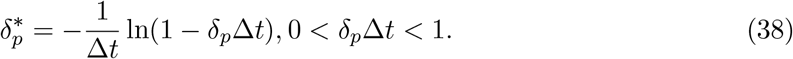

The multiplication rate is 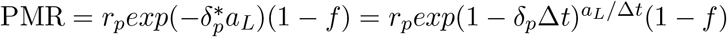.

**S1 Fig.**
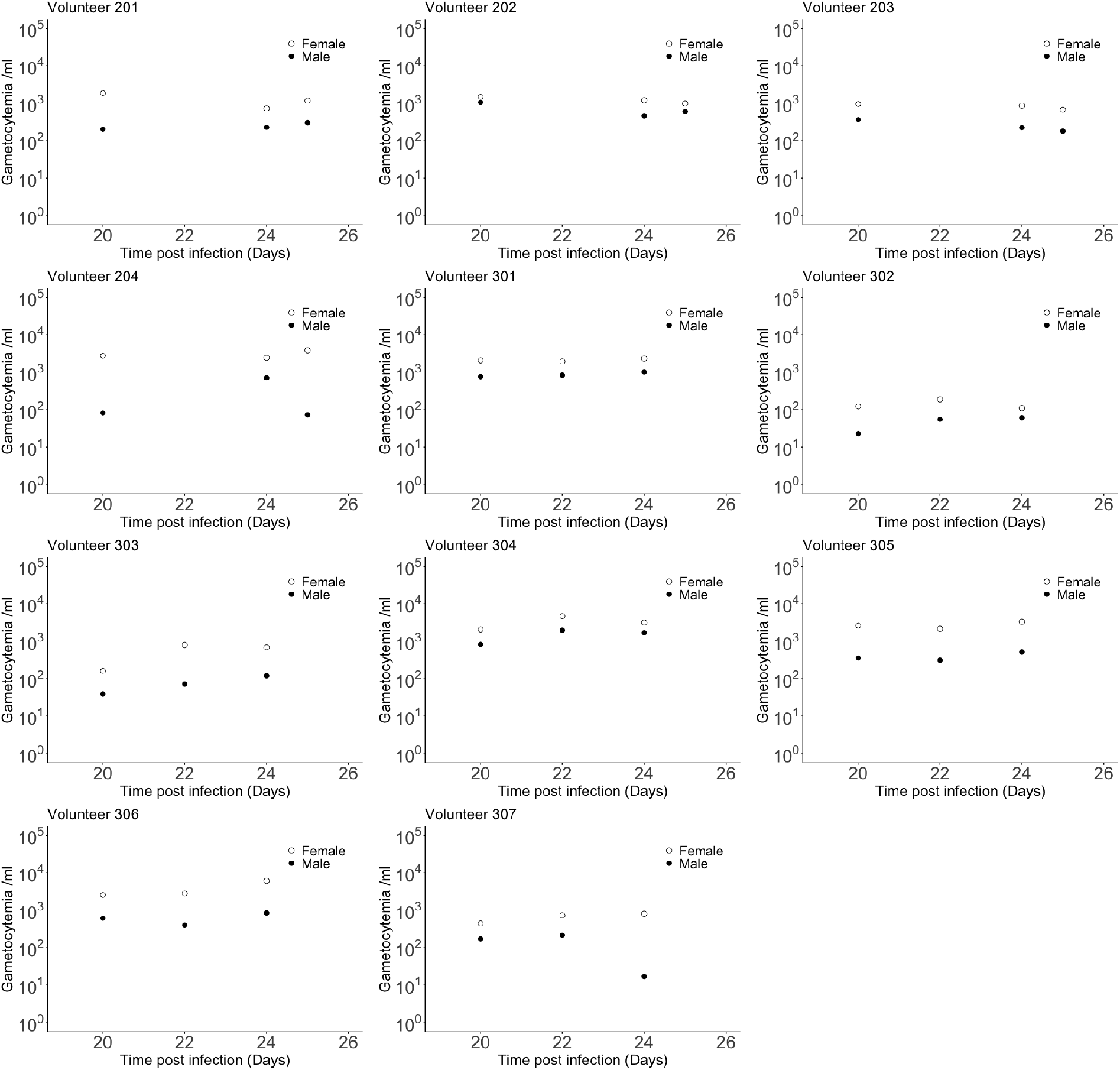
Direct feeding assay data including male (dots) and female (circles) gametocytemia for all 11 volunteers.

**S2 Fig.**
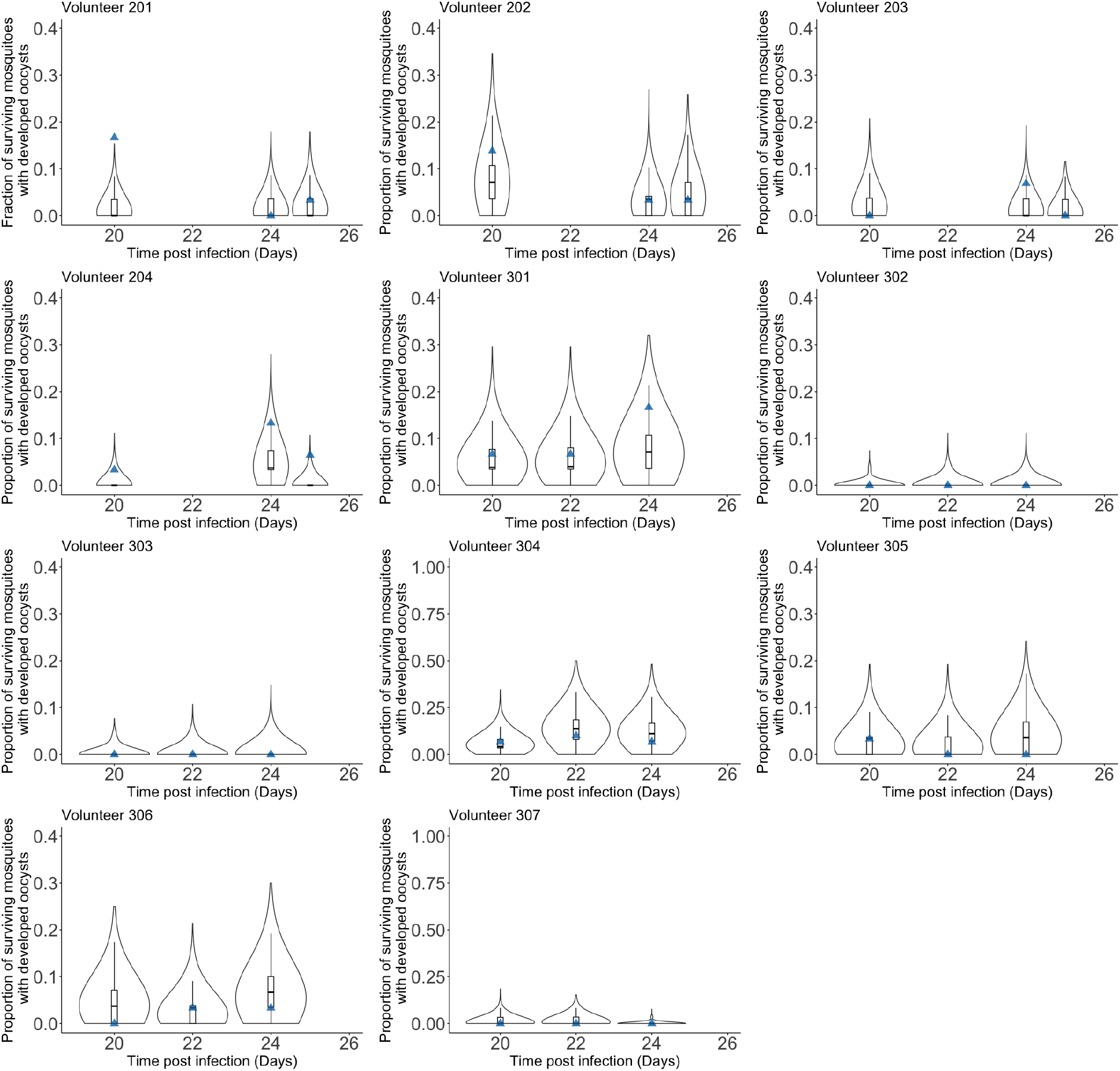
Results of model fitting for all 11 volunteers. Data are presented by blue triangles. The violin plots show the 5000 predictions of fraction of surviving mosquitoes with developed oocysts generated by 5000 posterior samples. Box plots show the 25%, 50% (median) and 75% quantiles.

**S3 Fig.**
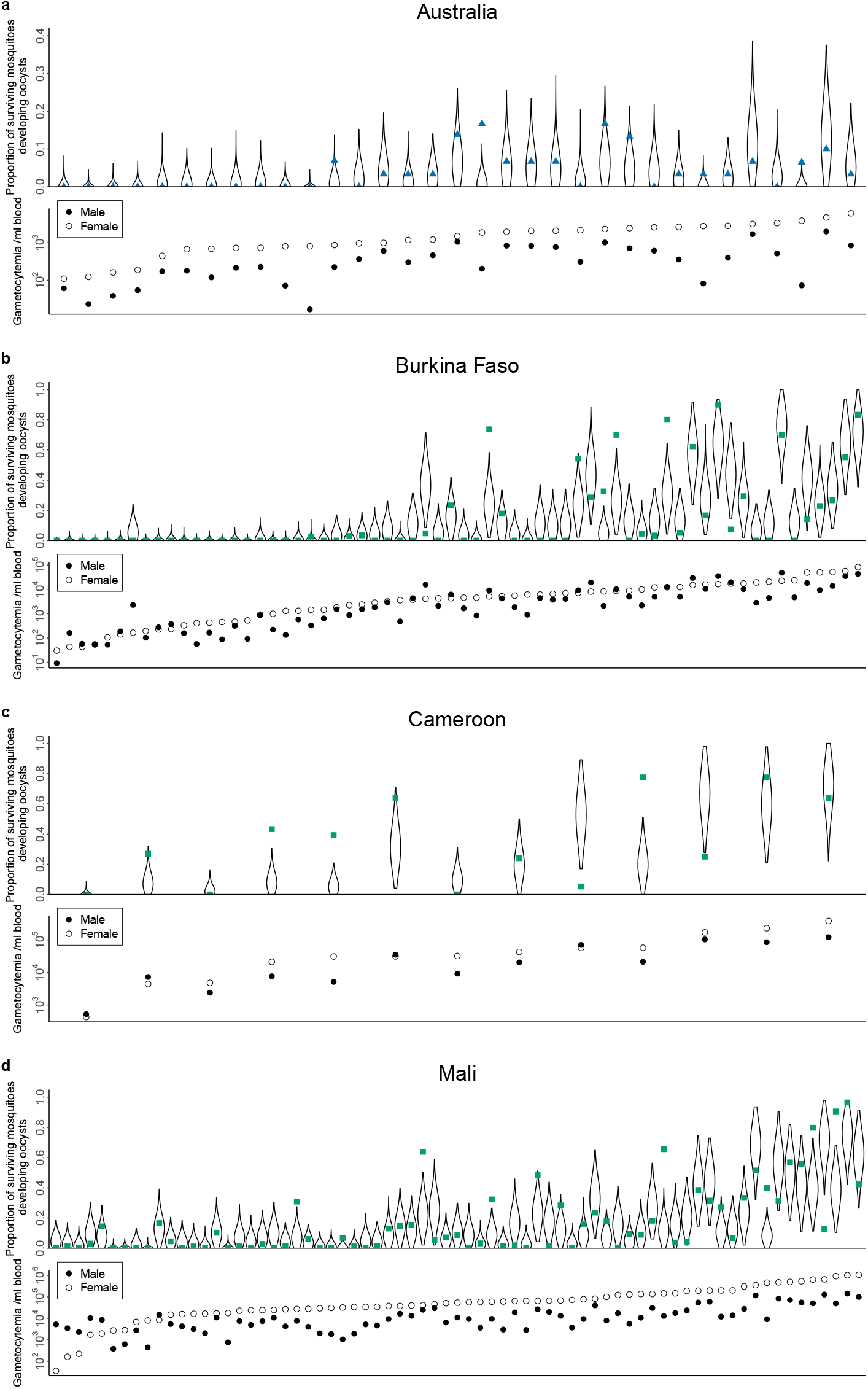
The model posterior estimates of the proportion of surviving mosquitoes developing oocysts ordered by female gametocytemia.

**S4 Fig.**
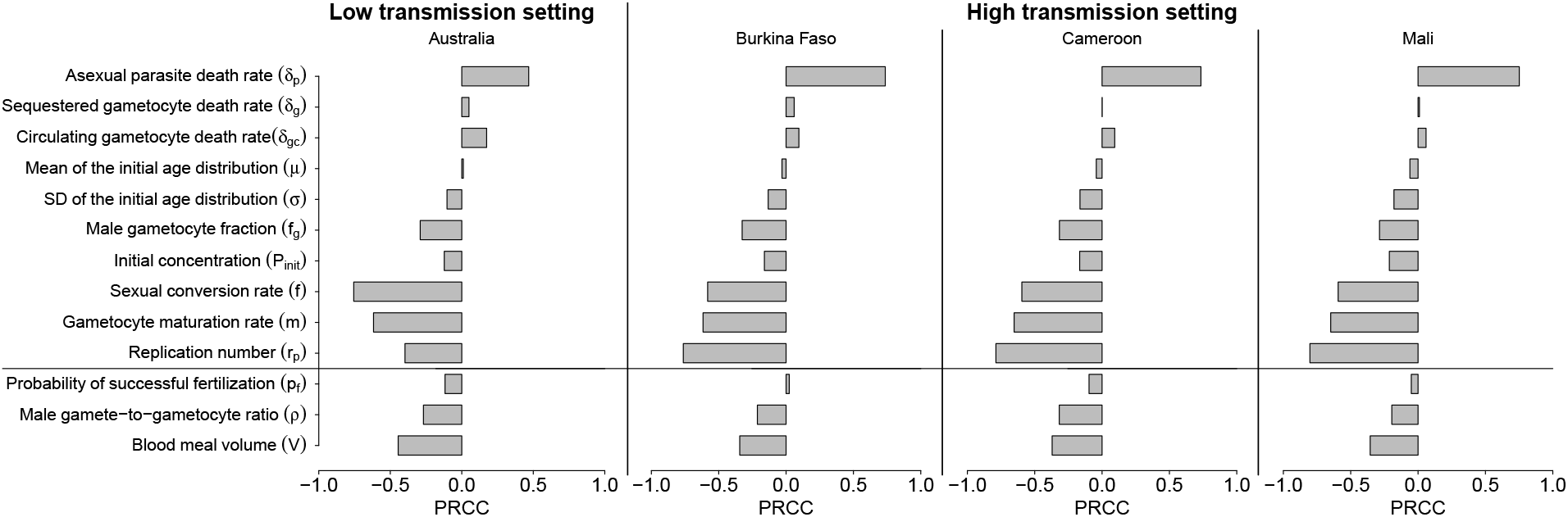
PRCC results between parameters and *t*_0.0005_.

**S5 Fig.**
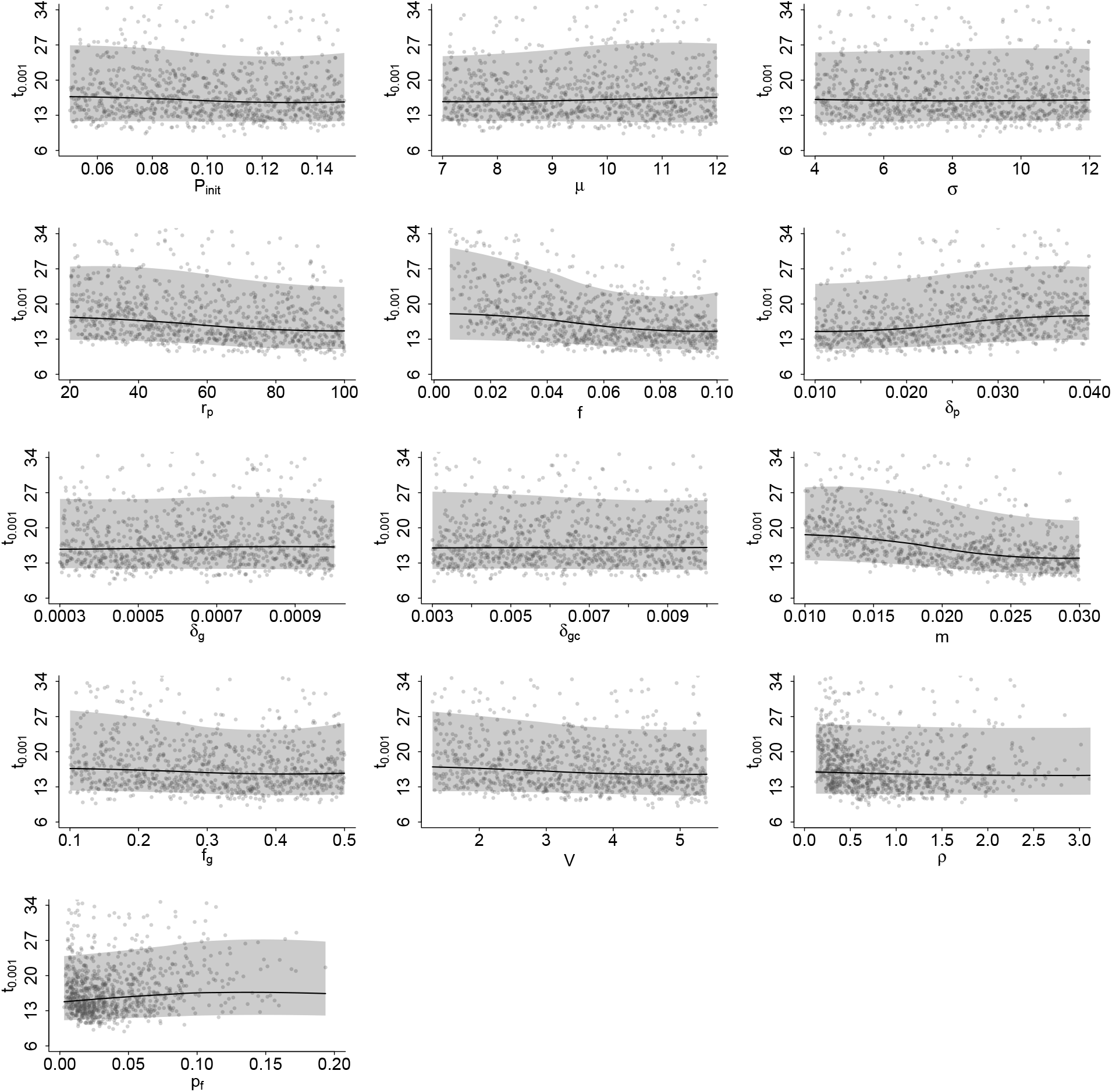
Marginal relationships between the sampled parameters and *t*_0.001_ for Australia. The black curves represents the local median relationships, and the grey shaded areas represent the corresponding 95% intervals. To calculate the median and 95% prediction intervals, the parameter range used in the PRCC analysis was divided into 500 equally spaced points. At each point, the local median and the 2.5th and 97.5th percentiles were calculated from the 50 nearest sampled parameter values.

**S6 Fig.**
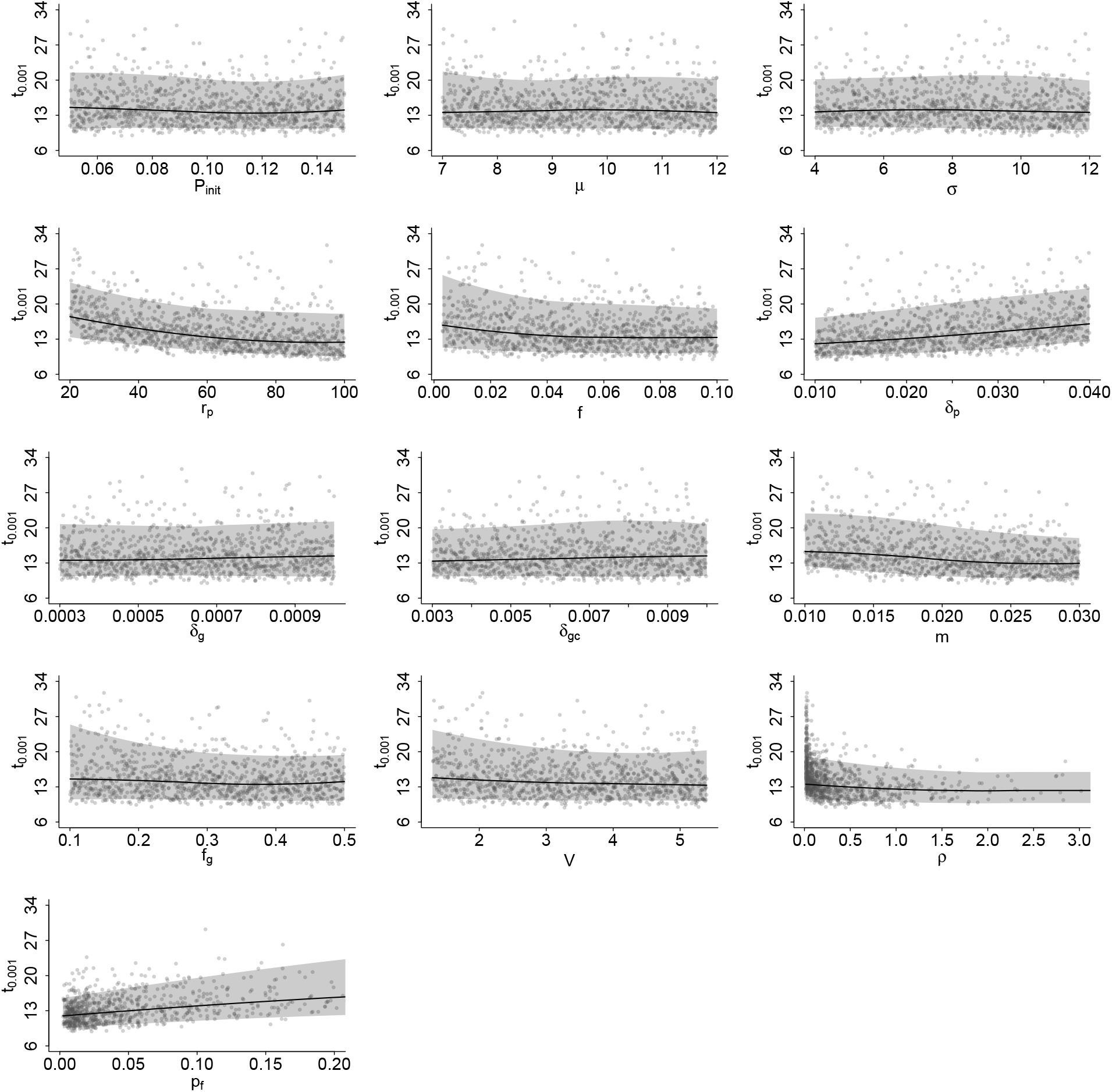
Marginal relationships between the sampled parameters and *t*_0.001_ for Burkina Faso. The black curves represents the local median relationships, and the grey shaded areas represent the corresponding 95% intervals. To calculate the median and 95% prediction intervals, the parameter range used in the PRCC analysis was divided into 500 equally spaced points. At each point, the local median and the 2.5th and 97.5th percentiles were calculated from the 50 nearest sampled parameter values.

**S7 Fig.**
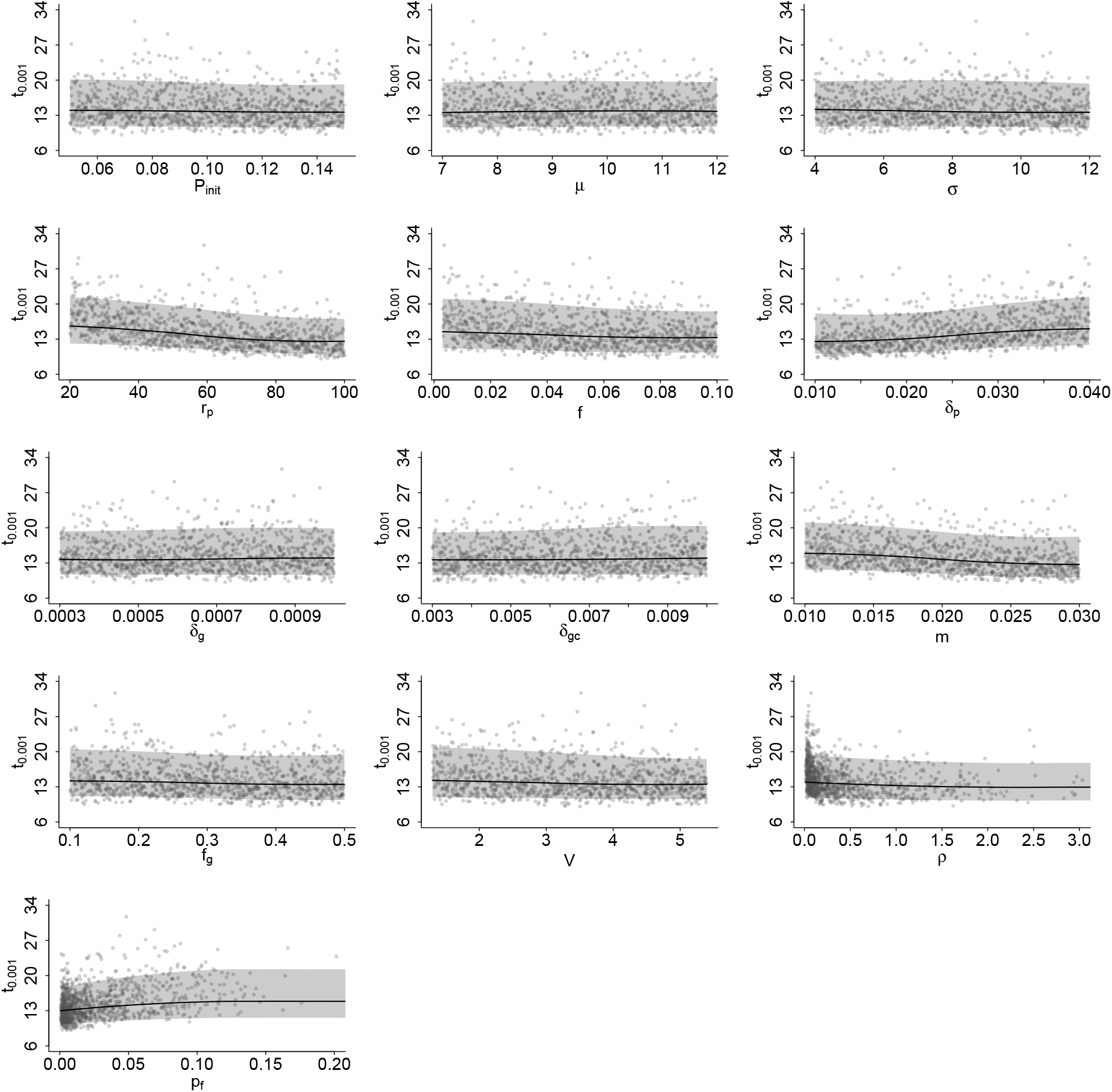
Marginal relationships between the sampled parameters and *t*_0.001_ for Cameroon. The black curves represents the local median relationships, and the grey shaded areas represent the corresponding 95% intervals. To calculate the median and 95% prediction intervals, the parameter range used in the PRCC analysis was divided into 500 equally spaced points. At each point, the local median and the 2.5th and 97.5th percentiles were calculated from the 50 nearest sampled parameter values.

**S8 Fig.**
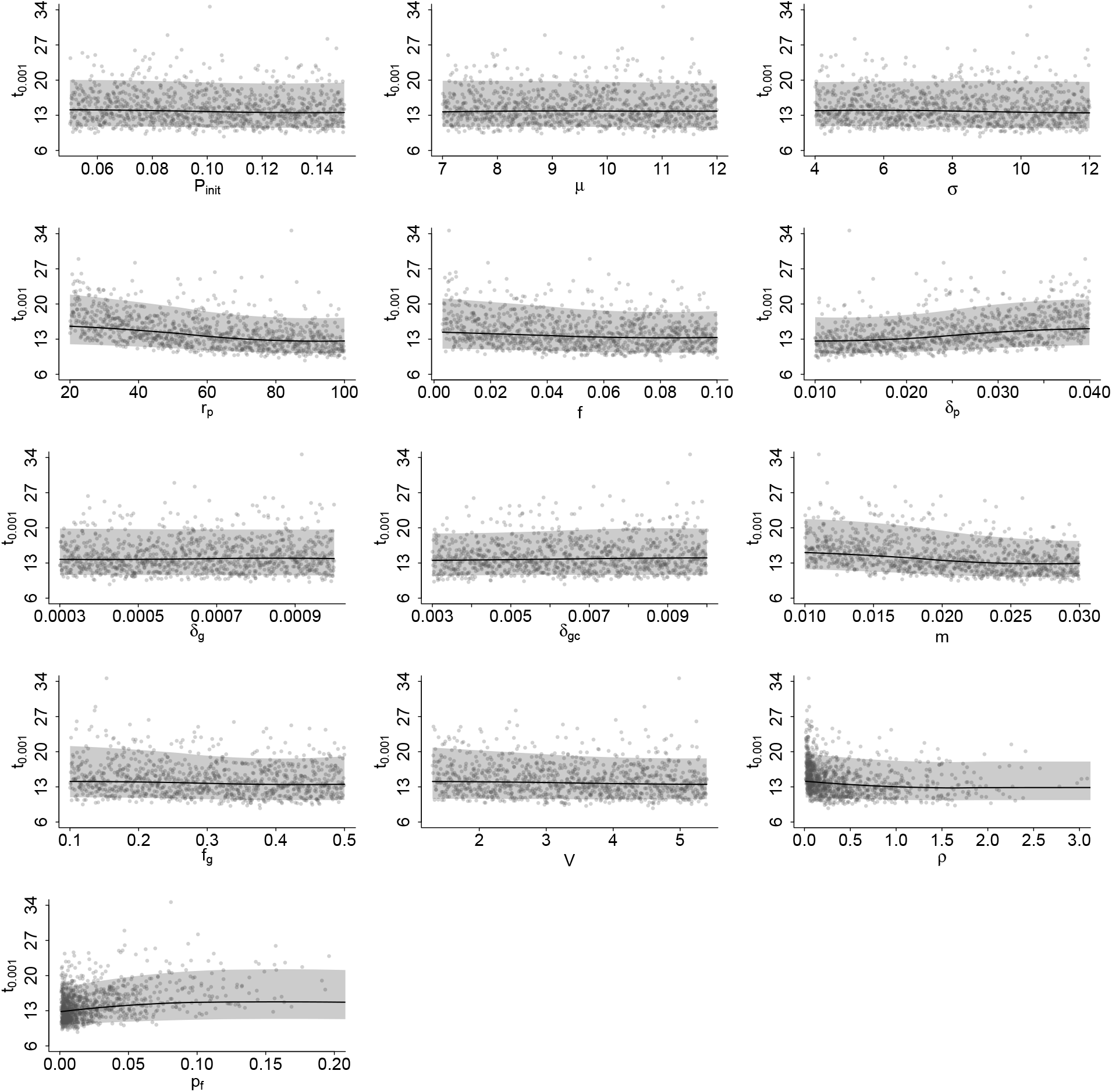
Marginal relationships between the sampled parameters and *t*_0.001_ for Mali. The black curves represents the local median relationships, and the grey shaded areas represent the corresponding 95% intervals. To calculate the median and 95% prediction intervals, the parameter range used in the PRCC analysis was divided into 500 equally spaced points. At each point, the local median and the 2.5th and 97.5th percentiles were calculated from the 50 nearest sampled parameter values.

**S9 Fig.**
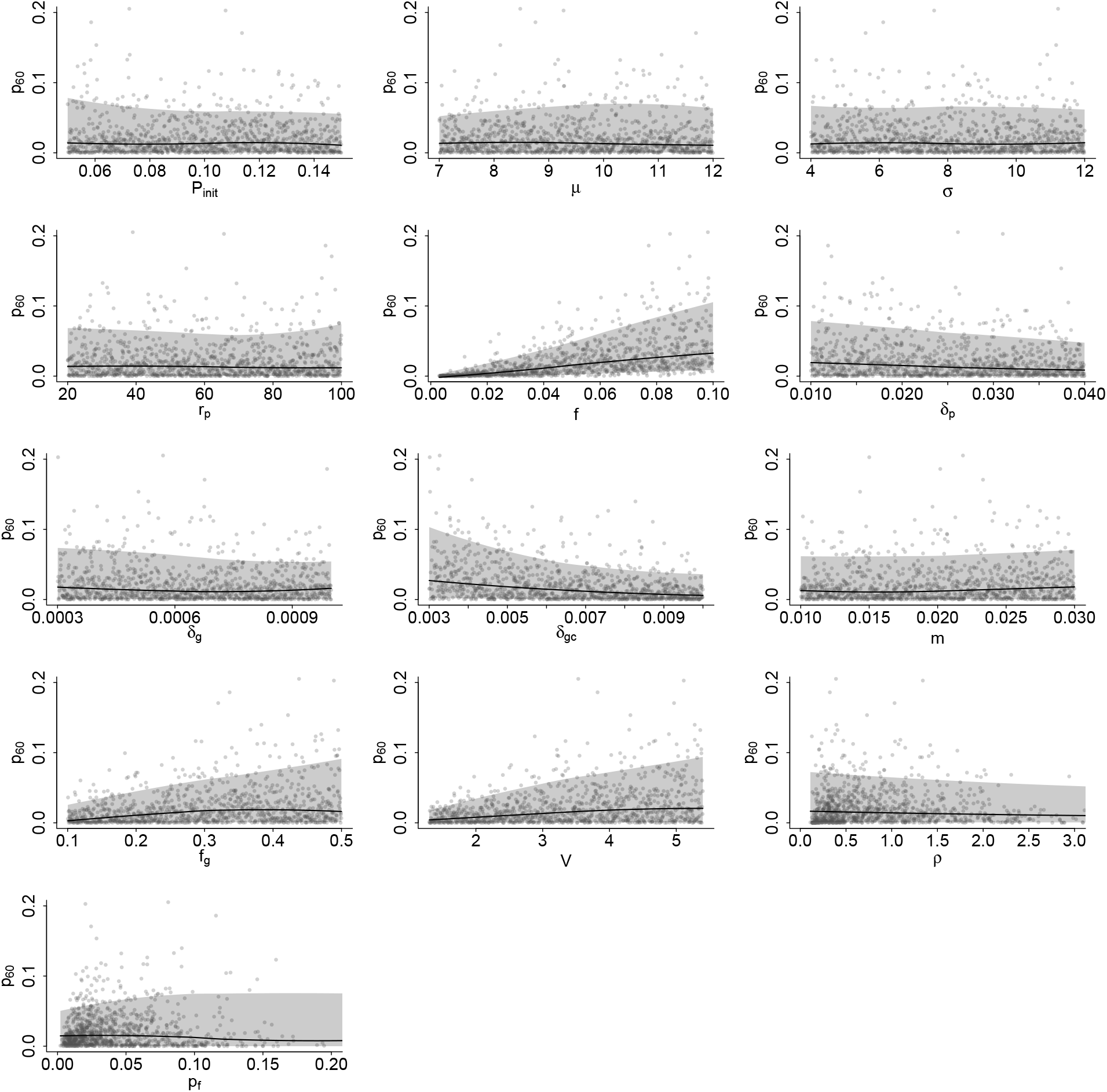
Marginal relationships between the sampled parameters and *p*_60_ for Australia. The black curves represents the local median relationships, and the grey shaded areas represent the corresponding 95% intervals. To calculate the median and 95% prediction intervals, the parameter range used in the PRCC analysis was divided into 500 equally spaced points. At each point, the local median and the 2.5th and 97.5th percentiles were calculated from the 50 nearest sampled parameter values.

**S10 Fig.**
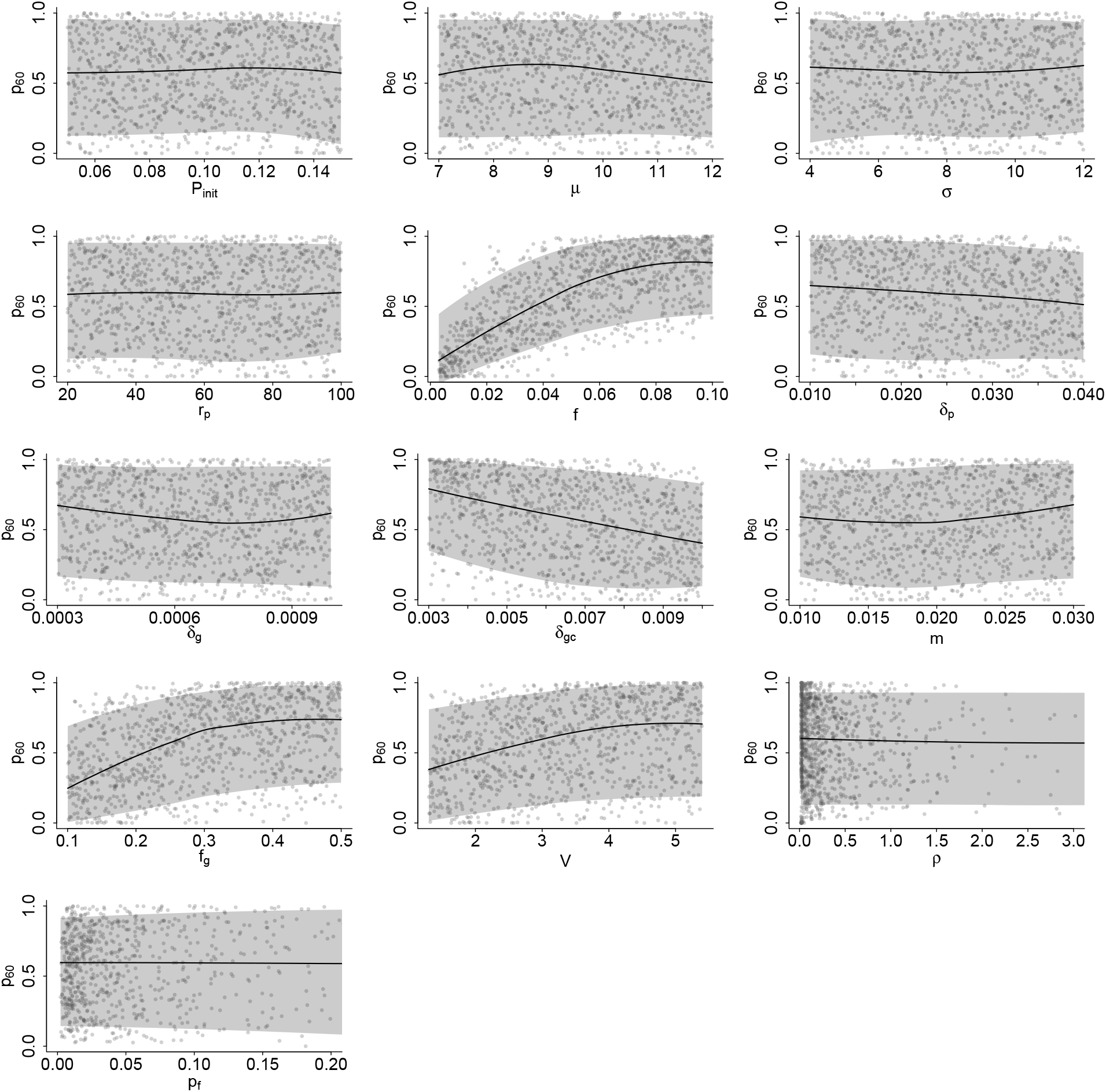
Marginal relationships between the sampled parameters and *p*_60_ for Burkina Faso. The black curves represents the local median relationships, and the grey shaded areas represent the corresponding 95% intervals. To calculate the median and 95% prediction intervals, the parameter range used in the PRCC analysis was divided into 500 equally spaced points. At each point, the local median and the 2.5th and 97.5th percentiles were calculated from the 50 nearest sampled parameter values.

**S11 Fig.**
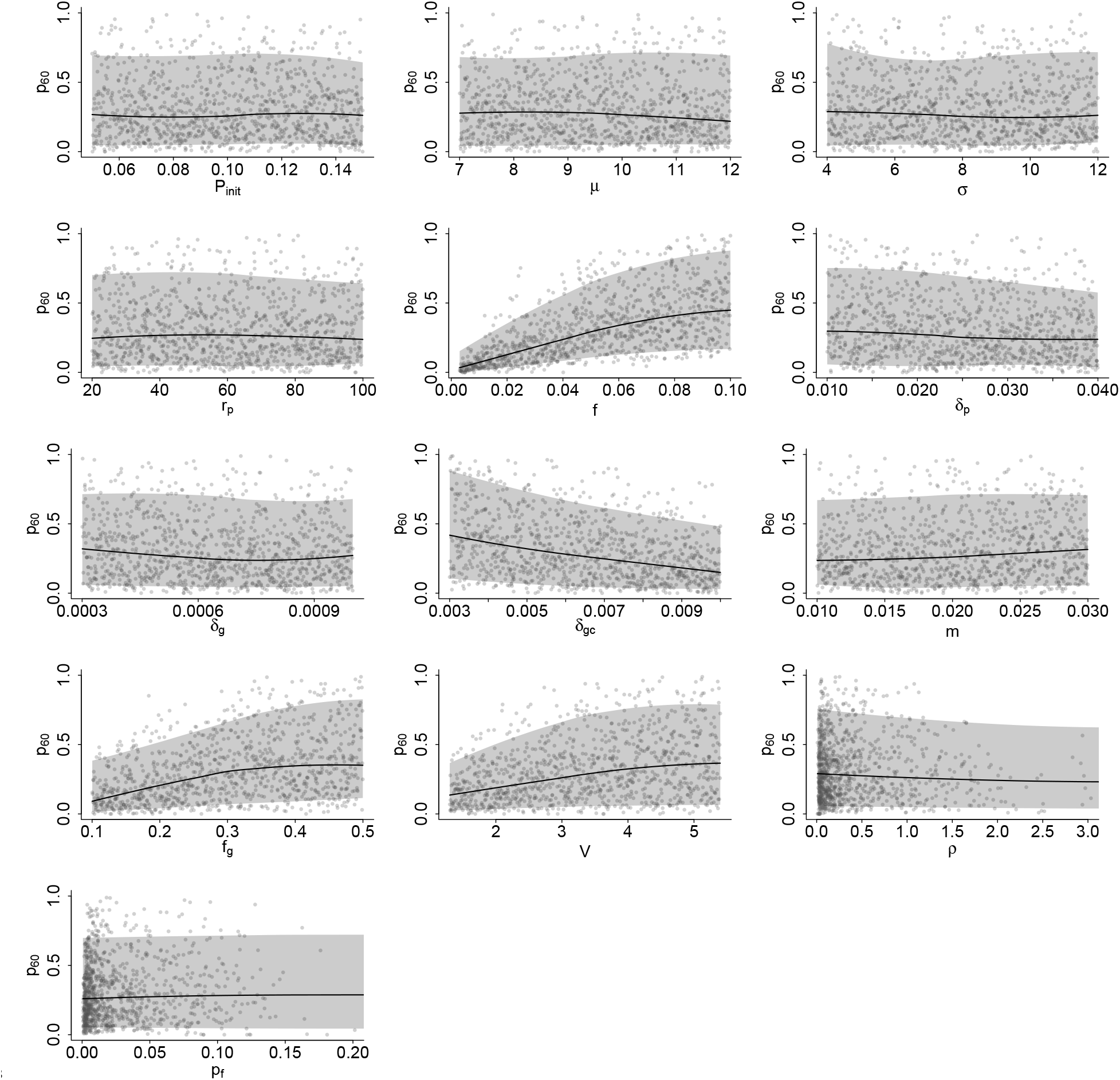
Marginal relationships between the sampled parameters and *p*_60_ for Cameroon. The black curves represents the local median relationships, and the grey shaded areas represent the corresponding 95% intervals. To calculate the median and 95% prediction intervals, the parameter range used in the PRCC analysis was divided into 500 equally spaced points. At each point, the local median and the 2.5th and 97.5th percentiles were calculated from the 50 nearest sampled parameter values.

**S12 Fig.**
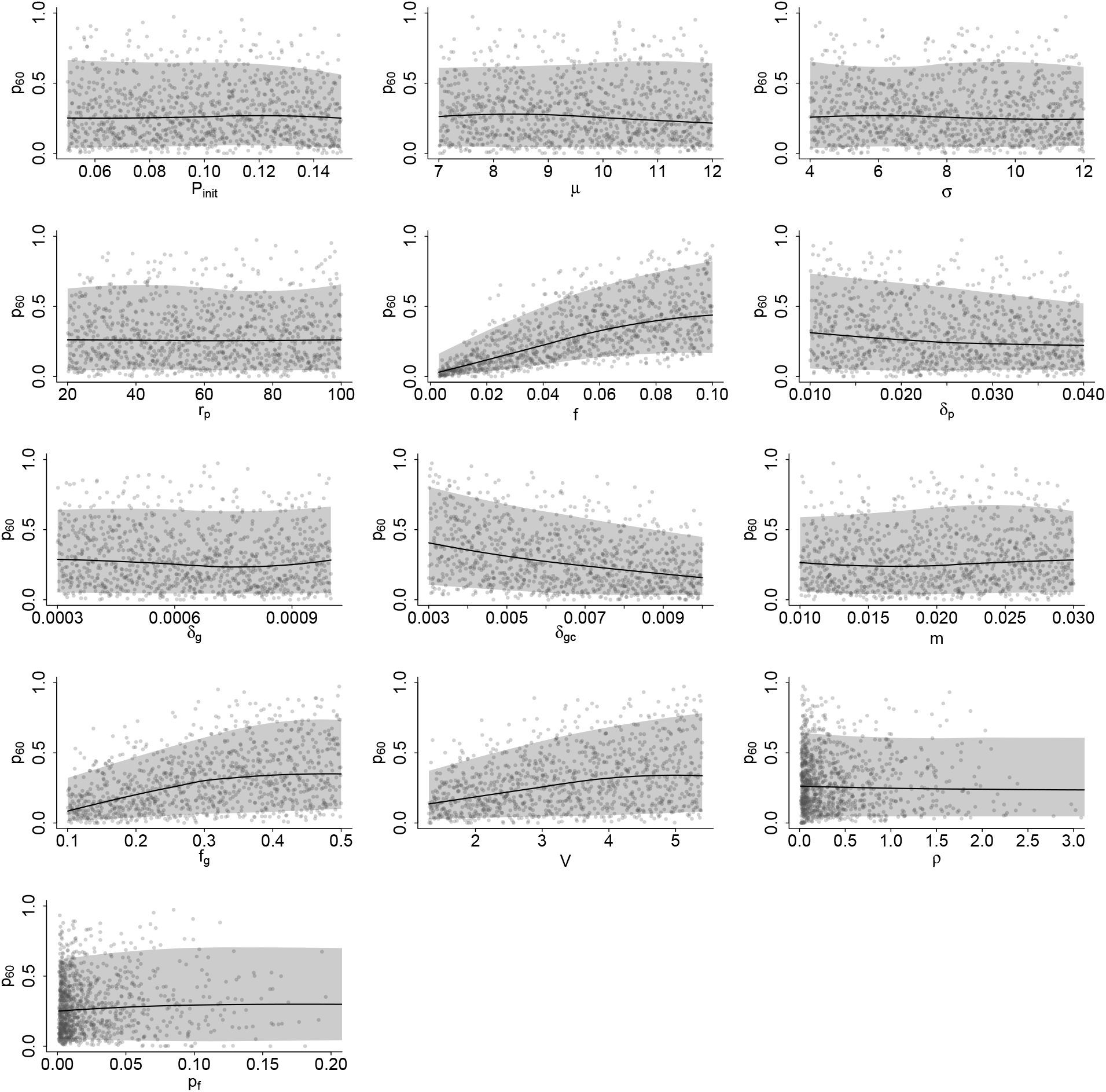
Marginal relationships between the sampled parameters and *p*_60_ for Mali. The black curves represents the local median relationships, and the grey shaded areas represent the corresponding 95% intervals. To calculate the median and 95% prediction intervals, the parameter range used in the PRCC analysis was divided into 500 equally spaced points. At each point, the local median and the 2.5th and 97.5th percentiles were calculated from the 50 nearest sampled parameter values.

**S13 Fig.**
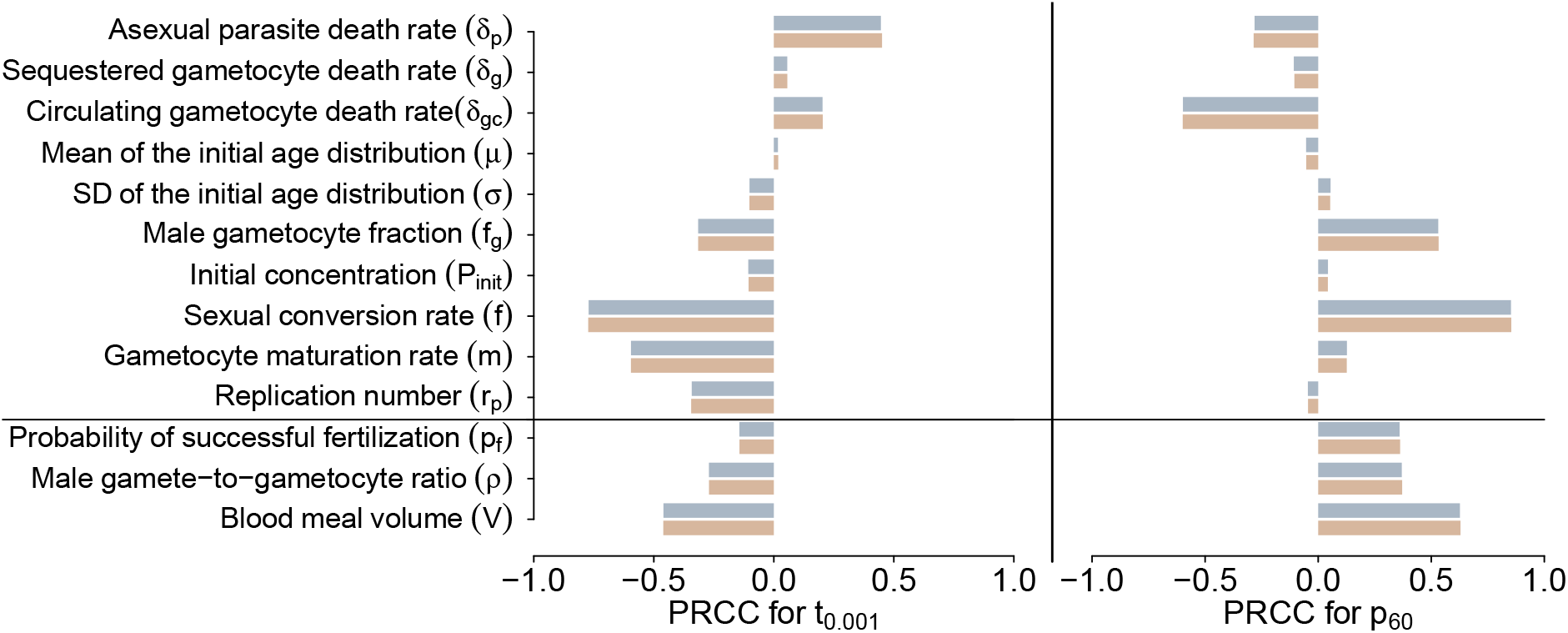
PRCC results for *t*_0.001_ and *p*_60_ under low transmission setting. Orange bars represent the PRCC values calculated using a single replicate, while blue bars represent the PRCC values calculated using the average of 100 replicates.

**S14 Fig.**
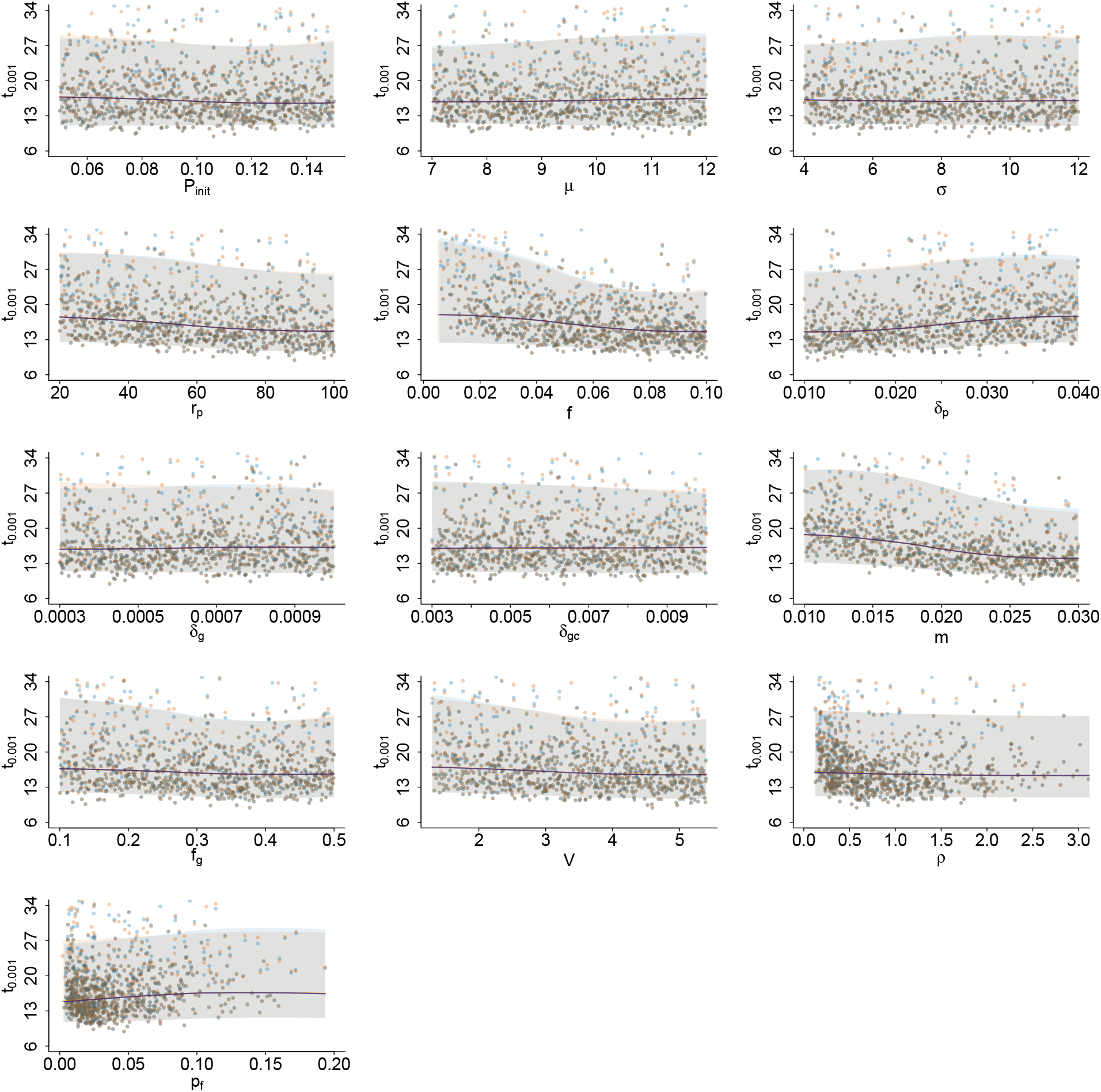
Marginal relationships between the sampled parameters and *t*_0.001_ under low transmission setting. The orange and blue dots show the *t*_0.001_ for the sampled parameter sets obtained using one replicate and the average of 100 replicates, respectively. The orange curves represent the local median relationships obtained using one replicate, and the orange shaded areas represent the corresponding 95% prediction intervals. The blue curves represent the local median relationships based on the average of 100 replicates, and the blue shaded areas represent the corresponding 95% prediction intervals. To calculate the median and 95% prediction intervals, the parameter range used in the PRCC analysis was divided into 500 equally spaced points. At each point, the local median and the 2.5th and 97.5th percentiles were calculated from the 50 nearest sampled parameter values.

**S15 Fig.**
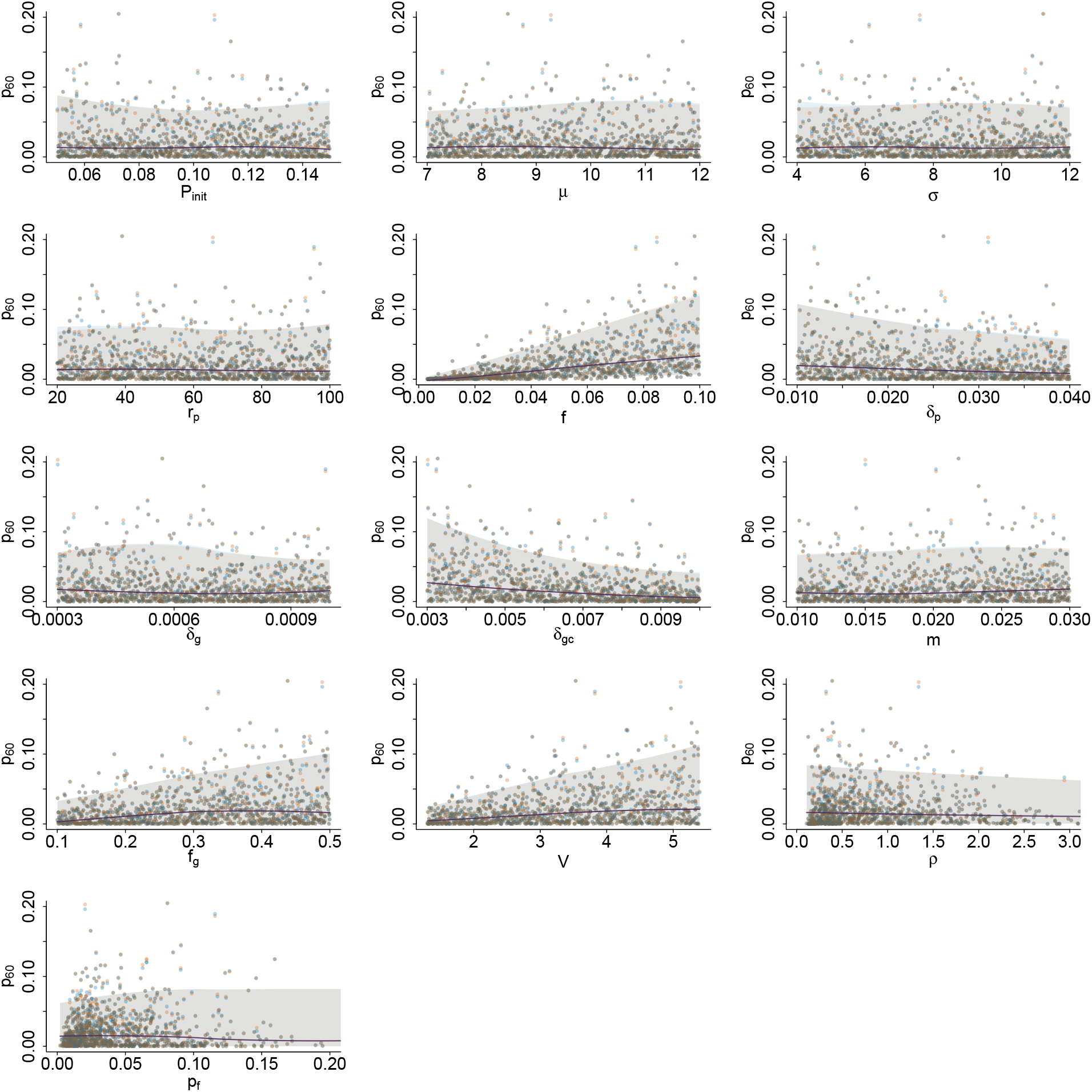
Marginal relationships between the sampled parameters and *p*_60_ under low transmission setting. The orange and blue dots show the *p*_60_ for the sampled parameter sets obtained using one replicate and the average of 100 replicates, respectively. The orange curves represent the local median relationships obtained using one replicate, and the orange shaded areas represent the corresponding 95% prediction intervals. The blue curves represent the local median relationships based on the average of 100 replicates, and the blue shaded areas represent the corresponding 95% prediction intervals. To calculate the median and 95% prediction intervals, the parameter range used in the PRCC analysis was divided into 500 equally spaced points. At each point, the local median and the 2.5th and 97.5th percentiles were calculated from the 50 nearest sampled parameter values.

**S16 Fig.**
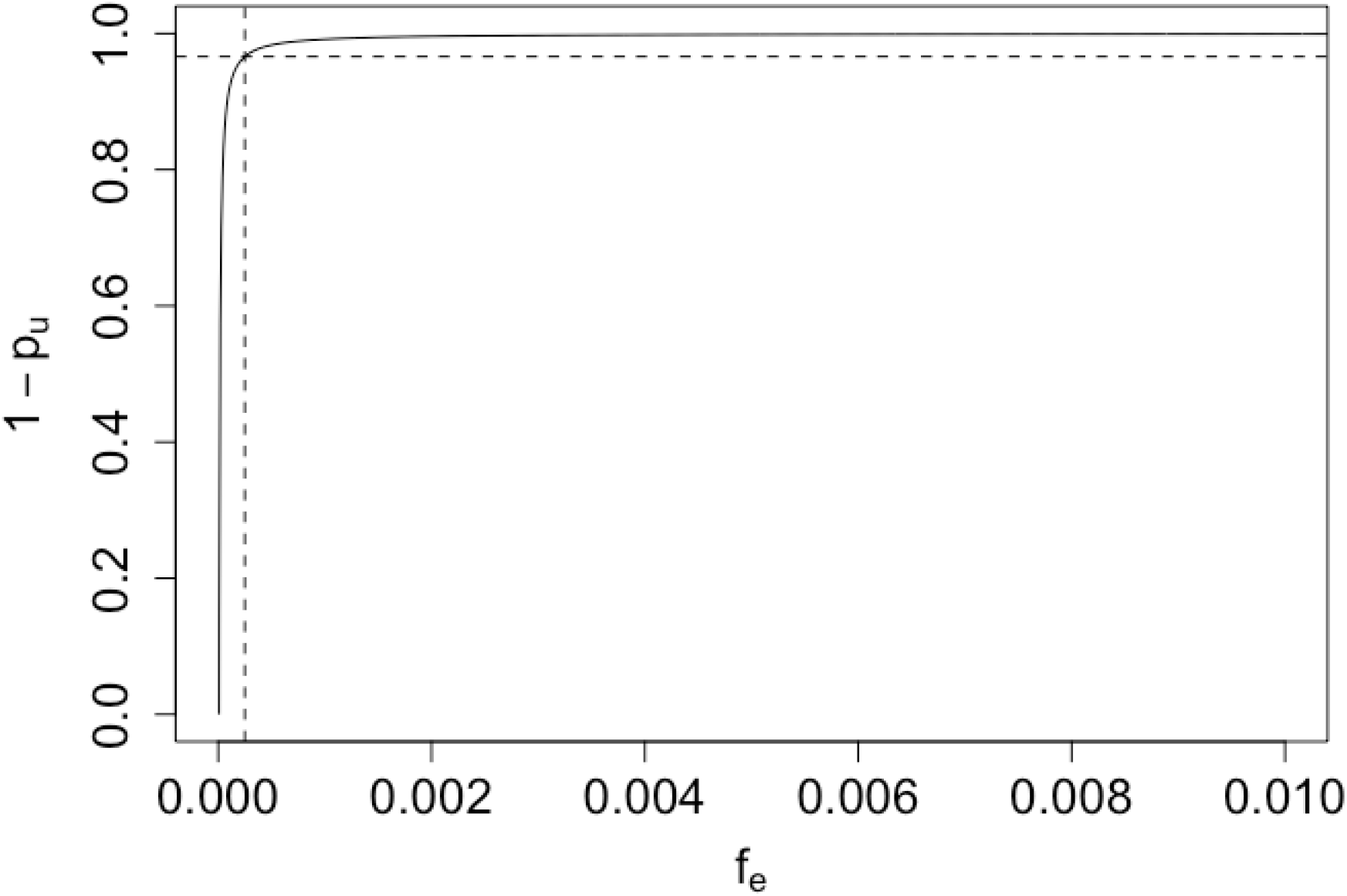
Relationship between *f*_*e*_ and 1 − *p*_*u*_. The plot illustrates how changes in *f*_*e*_ influence the variance of the binomial distributions determining the number of male and female gametocytes taken by a mosquito during a blood meal, which is given by 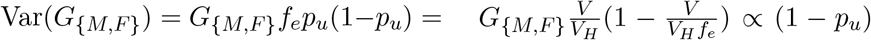. The dashed lines represent the benchmark value of *f*_*e*_ and corresponding 1 − *p*_*u*_.

